# SLC25A26-mediated SAM compartmentalization coordinates translation and bioenergetics during cardiac hypertrophy

**DOI:** 10.1101/2024.07.30.24311193

**Authors:** Ningning Guo, Jian Lv, Yu Fang, Qiuxiao Guo, Jiajie Li, Junmei Wang, Xiao Ma, Qingqing Yan, Fuqing Jiang, Shuiyun Wang, Li Wang, Zhihua Wang

## Abstract

**BACKGROUND:** The heart undergoes hypertrophy as a compensatory mechanism to cope with increased hemodynamic stress, and it can transition into a primary driver of heart failure. Pathological cardiac hypertrophy is characterized by excess protein synthesis. Protein translation is an energy-intensive process that necessitates an inherent mechanism to flexibly fine-tune intracellular bioenergetics according to the translation status; however, such a molecular link remains lacking.

**METHODS:** *Slc25a26* knockout and cardiac-specific conditional knockout mouse models were generated to explore its function *in vivo*. Reconstructed adeno-associated virus was used to overexpress *Slc25a26 in vivo*. Cardiac hypertrophy was established by transaortic constriction (TAC) surgery. Neonatal rat ventricular myocytes were isolated and cultured to evaluate the role of SLC25A26 in cardiomyocyte growth and mitochondrial biology *in vitro*. RNA sequencing was conducted to explore the regulatory mechanism by SLC25A26. m^1^A-modified tRNAs were profiled by RNA immuno-precipitation sequencing. Label-free proteomics was performed to profile the nascent peptides affected by S-adenosylmethionine (SAM).

**RESULTS:** We show that cardiomyocytes are among the top cell types expressing the SAM transporter SLC25A26, which maintains low-level cytoplasmic SAM in the heart. SAM biosynthesis is activated during cardiac hypertrophy, and feedforwardly mobilizes the mitochondrial translocation of SLC25A26 to shuttle excessive SAM into mitochondria. Systemic deletion of *Slc25a26* causes embryonic lethality. Cardiac-specific deletion of *Slc25a26* causes spontaneous heart failure and exacerbates cardiac hypertrophy induced by transaortic constriction. SLC25A26 overexpression, both before or after TAC surgery, rescues the hypertrophic pathologies and protects from heart failure. Mechanistically, SLC25A26 maintains low-level cytoplasmic SAM to restrict tRNA m^1^A modifications, particularly at A58 and A75, therefore decelerating translation initiation and modulating tRNA usage. Simultaneously, SLC25A26-mediated SAM accumulation in mitochondria maintains mitochondrial fitness for optimal energy production.

**CONCLUSIONS:** These findings reveal a previously unrecognized role of SLC25A26-mediated SAM compartmentalization in synchronizing translation and bioenergetics. Targeting intracellular SAM distribution would be a promising therapeutic strategy to treat cardiac hypertrophy and heart failure.

**Clinical Perspective:** *What Is New?:*

- An activation of S-adenosylmethionine (SAM) biosynthesis during cardiac hypertrophy boosts a feed-forward mitochondrial translocation of its transporter SLC25A26 to shuttle excessive SAM into mitochondria.
- SLC25A26-mediated cytoplasmic SAM containment restricts translation through inhibiting TRMT61A-mediated tRNA m^1^A modifications, particularly at A58 and A75, which modulates translation initiation and codon usage.
- SLC25A26-mediated mitochondrial SAM accumulation enhances mtDNA methylation and is required for the implement of mitochondrial fission and mitophagy, therefore maintaining optimal bioenergetics.
- Cardiac-specific knockout of *Slc25a26* causes spontaneous heart failure, and exacerbates transaortic constriction (TAC)-induced cardiac hypertrophy, while its overexpression rescues the hypertrophic pathologies.

*What Are the Clinical Implications?:*

- Cardiomyocyte-specific expression of SLC25A26 maintains low-level cytoplasmic SAM and contributes to the relatively low protein synthesis rate in the heart.
- Targeting intracellular SAM distribution would be a promising therapeutic strategy to treat cardiac hypertrophy and heart failure.

## INTRODUCTION

Cardiomyocytes predominantly express myofilament-related proteins, which constitute more than half of the entire translatome.^1^ The contraction and relaxation of the myofilament require substantial energy to maintain optimal cardiac performance.^2, 3^ Notably, translation also consumes a significant portion of the cellular energy expenditure *per se*.^4–6^ The translation activity is stringently monitored to ensure appropriate justification of the energetic investment.^6^ Thus, the interplay between contractility, translation and bioenergetics forms a complex triadic entanglement in working cardiomyocytes. In theory, there should be an inherent mechanism that coordinates these processes to provide cardiomyocytes with the necessary adaptability under pathophysiological conditions; however, such a molecular link is still lacking.^7, 8^

The heart undergoes hypertrophy as a compensatory mechanism to cope with increased hemodynamic stress; however, if the stress persists without timely relief, hypertrophy can transition into a primary driver of heart failure.^9^ Pathological cardiac hypertrophy is characterized by enlarged cardiomyocyte size and excess protein synthesis.^10–12^ Despite early indications of increased ribosome biogenesis,^13–15^ little headway has been made in understanding the regulatory mechanisms governing translational control during cardiac hypertrophy.^16^ While gene abnormalities in cardiac hypertrophy have been extensively examined at the transcriptional level, it is important to note that the cardiac translatome cannot be precisely dictated by the transcriptome due to ribosome efficacy and mRNA selectivity.^17–20^

The pathological growth of cardiomyocytes is accompanied by an enhancement of translation initiation.^12^ Phosphorylation of eukaryotic translation initiation factor 2Bε by glycogen synthase kinase-3β contributes to the enhanced translation activity in β-adrenergic cardiac hypertrophy.^21^ A recent study revealed that activation of inositol-requiring enzyme 1α during cardiac hypertrophy facilitates the formation of the translation initiation complex around endoplasmic reticulum and promotes the translation of transcripts with 5’ terminal oligopyrimidine motifs.^22^ Translation starts with the relief of the inhibitor eukaryotic translation initiation factor 4E binding protein 1 (4E-BP1) through mechanistic target of rapamycin kinase complex 1 (mTORC1)-mediated phosphorylation, a process that is required for the hypertrophic growth of the heart.^23, 24^ However, cardiac-specific deletion of *Raptor*, the core component of mTORC1,^25^ impairs adaptive hypertrophy, but causes heart failure in mice.^26^ Deleting p70 ribosomal S6 kinase, another target of mTORC1 involving in translational activation, has no effect on pressure-overload-induced cardiac hypertrophy.^27^

Regulations at the tRNA level play a crucial role in fine-tuning protein synthesis in response to various cellular conditions and signals.^28^ Modifications to tRNAs affect their stability, aminoacylation, and codon recognition properties, ultimately influencing the overall translation process.^29^ The initiator tRNA (tRNA^iMet^) can be methylated at the N(1) position of A58 (m^1^A58) by the tRNA methyltransferase (TRMT) 6/61A complex, leading to an enhancement of translation initiation.^30, 31^ Moreover, tRNA modifications can change the status of the tRNA pool, leading to biased codon usage and altered translating rate of individual mRNAs.^32^ However, it remain unclear whether tRNA modifications are involved in the translational control during cardiac hypertrophy.

The energetic cost of protein synthesis accounts for ∼30% of the total energy consumption in mammalian cells.^5^ In the heart, up to 95% of adenosine triphosphate (ATP) is derived from mitochondria, which occupy ∼30% of the total myocardial mass.^33^ Mitochondrial fitness is crucial for maintaining the normal heart function; and its disorder plays a causal role in heart diseases.^34–36^ Mitochondrial oxidative capacity has been reported to be either preserved or even enhanced in cardiac hypertrophy,^37^ but the regulatory mechanism of mitochondrial dynamics under hypertrophic stress remains to be elucidated.

S-adenosylmethionine (SAM) is the sole methyl donor for macromolecular methylation reactions.^38^ Its biosynthesis is catalyzed by methionine adenosyltransferase 2A (MAT2A) using methionine and ATP as the substrates.^39^ The mitochondrial shuttling of SAM requires a transporter named solute carrier family 25 member 26 (SLC25A26; also known as SAMC).^40^ We find that the cardiomyocytes express low-level MAT2A but high-level SLC25A26, making mitochondria a pool of the intracellular SAM. Whereas the cytoplasmic SAM (cytoSAM) enhances translation activity via tRNA m^1^A modification, the mitochondrial SAM (mitoSAM) maintains mitochondrial fitness through promoting fission and mitophagy. During cardiac hypertrophy, an increase in SAM biosynthesis boosts the mitochondrial translocation of SLC25A26 in a feedforward manner to transport excessive cytoSAM into mitochondria. Cardiac-specific deletion of *Slc25a26* in mice causes spontaneous heart failure and exacerbates cardiac hypertrophy induced by transaortic constriction, whereas its overexpression rescues the hypertrophic pathologies. Our findings demonstrate that subcellular distribution of SAM links bioenergetics with translation in cardiomyocytes, and represents a promising target for the treatment of cardiac hypertrophy and heart failure.

## METHODS

Collection and usage of human samples were approved by the Ethics Committee of the Fuwai Hospital Chinese Academy of Medical Sciences, Shenzhen [SP2023133(01)]. Animal experiments were reviewed and approved by the Institutional Animal Care and Use Committee (IACUC) of Renmin Hospital of Wuhan University (No. 20180508) and the IACUC of Fuwai Hospital Chinese Academy of Medical Sciences, Shenzhen (SP2023059). More details of the experimental procedures are included in the Supplemental Material.

### Data Availability

RNA-seq data have been deposited in NCBI’s Gene Expression Omnibus (GEO) repository (accession: GSE254565 and GSE173737). Online microarray datasets are available from NCBI’s GEO repository with corresponding accession codes as described in details in Supplemental Methods. Any additional information reported in this paper is available from the lead contact upon request.

### Statistical Analyses

Statistical analyses were performed using GraphPad Prism 8.0 and R software. All experimental data were presented as mean ± SEM of at least three independent experiments, unless denoted elsewhere. Statistical significance for multiple comparisons was determined by one-way ANOVA or two-way ANOVA followed by Tukey’s test. Bonferroni adjustment was used for *post hoc* analysis. Student’s *t* test was used for comparisons between two groups. *P* < 0.05 was considered statistically significant.

## RESULTS

### Feedforward Mitochondrial SAM Transport by SLC25A26 during Cardiac Hypertrophy

SAM is mainly synthesized by MAT2A in cytosol,^41^ and can be shuttled into mitochondria by SLC25A26 before donating its methyl group (Figure 1A). Human single-cell sequencing data highlighted skeletal myocytes as the cell type with the highest *MAT2A* expression, versus cardiomyocytes to be the lowest one (Figure 1B). On the contrary, cardiomyocytes were among the top cell types expressing *SLC25A26* (Figure 1C). Western blot analysis using mouse tissues validated the low expression of MAT2A in the heart, but it showed a high signal in the liver (Figure 1D), possibly due to the antibody misrecognition with its hepatic-specific homolog MAT1A (Figure S1A). Intriguingly, SLC25A26 was specifically enriched in the heart (Figure 1D), exclusively in cardiomyocytes (Figure S1B). Enzyme-linked immunosorbent assay (ELISA) showed that the level of cytoSAM was extremely low compared with mitoSAM when balanced to the protein concentration; and the level of mitoSAM in the heart was roughly 3-fold higher than that in the skeletal muscle and liver (Figure 1E). These results suggest a special SAM distribution pattern implicated in cardiac function.

**Figure 1.**
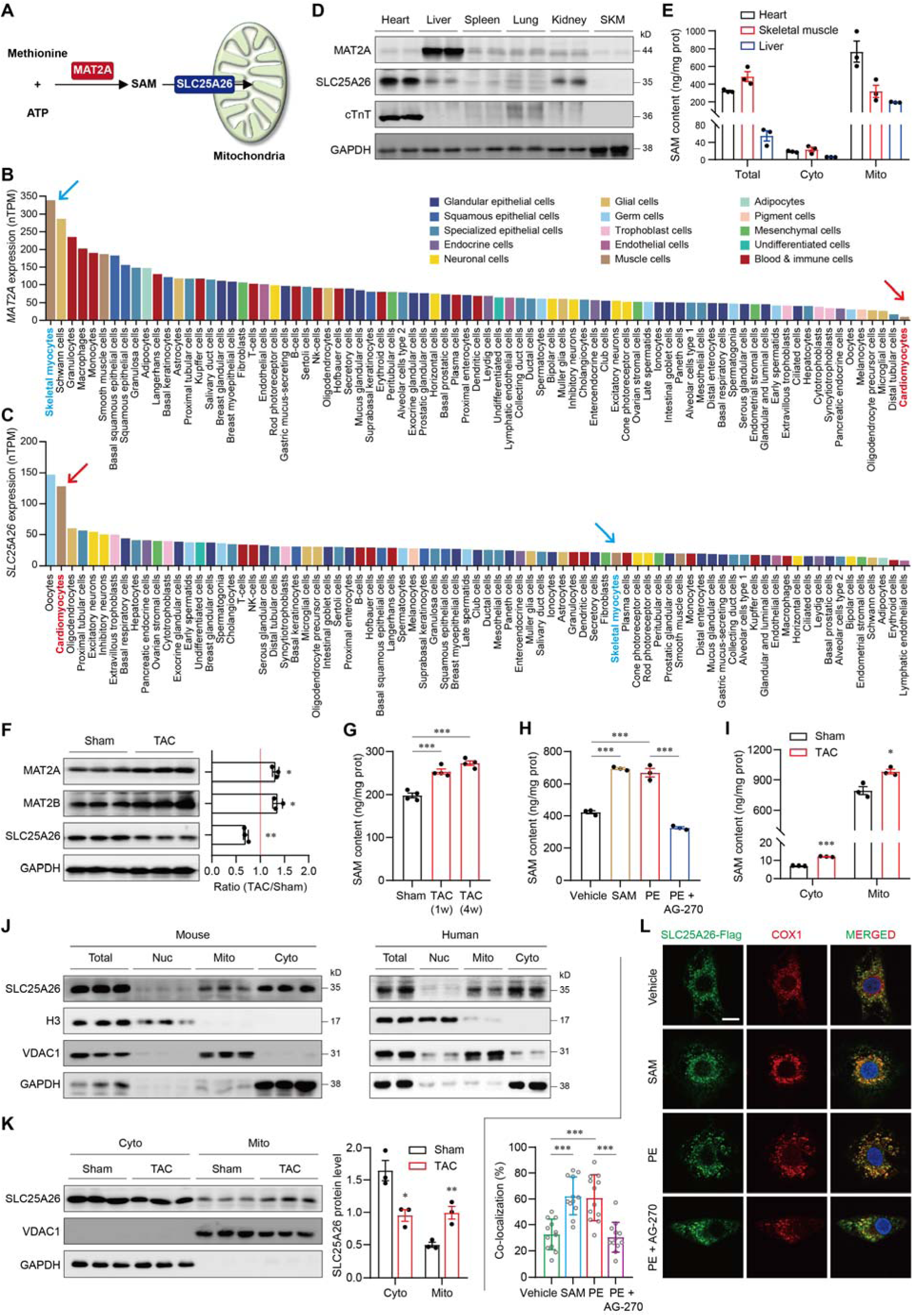
Feedforward SAM transport into mitochondria by SLC25A26 during cardiac hypertrophy. **A**, Schematic illustrating the SAM biosynthesis and mitochondrial transport pathways. **B** and **C**, Cell-type-specific expression of *MAT2A* (**B**) and *SLC25A26* (**C**) from a single-cell sequencing database (Human Protein Atlas; https://www.proteinatlas.org/). **D**, Western blot analysis showing the tissue specificity of MAT2A and SLC25A26 in mice (*N* = 2). Troponin T2, cardiac type (cTnT) was used as a positive heart control. Glyceraldehyde-3-phosphate dehydrogenase (GAPDH) was used as a loading controls. SKM, skeletal muscle. **E**, Total and subcellular SAM contents in the heart, skeletal muscle and liver. *N* = 3. Cyto, cytoplasm; Mito, mitochondria. **F**, Immunoblots (left) and quantification data (right) showing the expressions of MAT2A, MAT2B and SLC25A26 in transaortic constriction (TAC)-induced cardiac hypertrophy. *N* = 3. **G**, Time-course effects of TAC on the SAM content in the heart measured by enzyme-linked immunosorbent assay (ELISA). *N* = 4. **H**, SAM contents in neonatal rat ventricular myocytes (NRVMs) treated with SAM (1μM) or phenylephrine (PE; 50μM), in the presence or absence of an MAT2A inhibitor AG-270 (3μM). *N* = 3. **I**, Effect of TAC on subcellular SAM distribution in fractionated mouse hearts. *N* = 3. **J**, Subcellular SLC25A26 localization in fractionated mouse (left; *N* =3) and human (right; *N* =2) hearts with H3, VDAC1 and GAPDH as nuclear (Nuc), Mito and Cyto controls, respectively. **K**, Immunoblots (left) and quantification data (right) showing the impact of TAC on subcellular SLC25A26 localization. *N* = 3. **L**, Representative immunofluorescence images (upper) and quantification data (lower) showing the co-localization of the Flag-tagged SLC25A26 with mitochondria (COX1) in NRVMs treated with SAM or PE, with or without AG-270. \*\*\**P* < 0.001; *N* = 12. Scale, 5μm. Data are mean ± SEM. *p < 0.05, **p < 0.01, ***p < 0.001 vs. sham or vehicle, (**F**, **I** and **K**, unpaired Student’s t test; **G**, **H**, **L**, one-way ANOVA with Tukey’s multiple comparison test).

We then asked how the SAM biology would be shaped under cardiac stress. Both MAT2A and its regulatory subunit MAT2B were significantly upregulated during the development of cardiac hypertrophy after transaortic constriction (TAC) surgery in mice (Figure 1F), concurrent with a time-course increase in the SAM content (Figure 1G). In isolated neonatal rat ventricular myocytes (NRVMs), hypertrophic stimulation by phenylephrine (PE; 50μM) also increased the SAM content to a similar extent as exogenous SAM supplementation (1μM; Figure 1H). The PE-induced SAM increase was completely blocked by an MAT2A inhibitor AG-270 (3μM; Figure 1H),^42^ suggesting an activation of SAM biosynthesis during cardiac hypertrophy.

Nevertheless, the expression of SLC25A26 showed a significant decrease after TAC (Figure 1F), which was consistent with observations in human hearts with dilated cardiomyopathy and mouse TAC databases (Figures S1C and S1D). Despite a decrease in SLC25A26 expression, the level of mitoSAM was still significantly elevated by TAC (Figure 1I), indicating an alteration of SAM-transporting activity. To our surprise, albeit originally designated as a mitochondrial protein,^40^ significant amount of SLC25A26 was detected in the cytoplasmic fraction of both human and mouse hearts (Figure 1J). Yet TAC surgery caused substantial translocation of SLC25A26 from cytosol to mitochondria (Figure 1K). Consistently, PE treatment in NRVMs also promoted the mitochondrial translocation of SLC25A26, an effect that could be mimicked by supplementing SAM but blocked by AG-270 (Figure 1L). These data suggest that the increased SAM biosynthesis during cardiac hypertrophy promotes the mitochondrial translocation of SLC25A26, thereby shuttling excessive cytoSAM into mitochondria in a feed-forward manner.

### SLC25A26 Is Required for Development and Normal Heart Function

To investigate the physiological function of SLC25A26, we tried to generate an *Slc25a26*-knockout mouse line using CRISPR-cas9 (Figures S2A and S2B). However, homozygous knockout of *Slc25a26* caused embryonic lethality (Figure S2C), suggesting an essential role of SLC25A26 in development. This is consistent with previous observations that loss-of-function mutations of human *SLC25A26* resulted in neonatal mortality.^43–45^

We then developed an inducible cardiac-specific conditional knockout (cKO) mouse line by generating an *Slc25a26*-flox mouse line and hybridizing the mice with the αMHC-MerCreMer transgenic mice (Figure 2A), so that enabling controllable *Slc25a26* deletion in adult cardiomyocytes (Figure 2B).^46^ *Slc25a26* cKO did not influence the heart weight but significantly increased the lung weight twenty weeks after tamoxifen administration (Figures 2C and 2D), a characteristic symptom of heart failure. Echocardiography showed that *Slc25a26* deficiency led to enlarged left ventricular internal diameter (LVID) both in diastole and in systole and impaired the ejection fraction (EF) and fraction shortening (FS) (Figures 2E through 2G). Moreover, the *Slc25a26*-cKO hearts exhibited severe myocardial fibrosis (Figure 2H) and robust inductions of pathological markers, including natriuretic peptide A (*Nppa*), natriuretic peptide B (*Nppb*), and myosin heavy chain 7 (*Myh7*), compared with the *Slc25a26*-flox hearts (Figure 2I). These data suggest an essential role of SLC25A26 in the maintenance of normal heart morphology and function.

**Figure 2.**
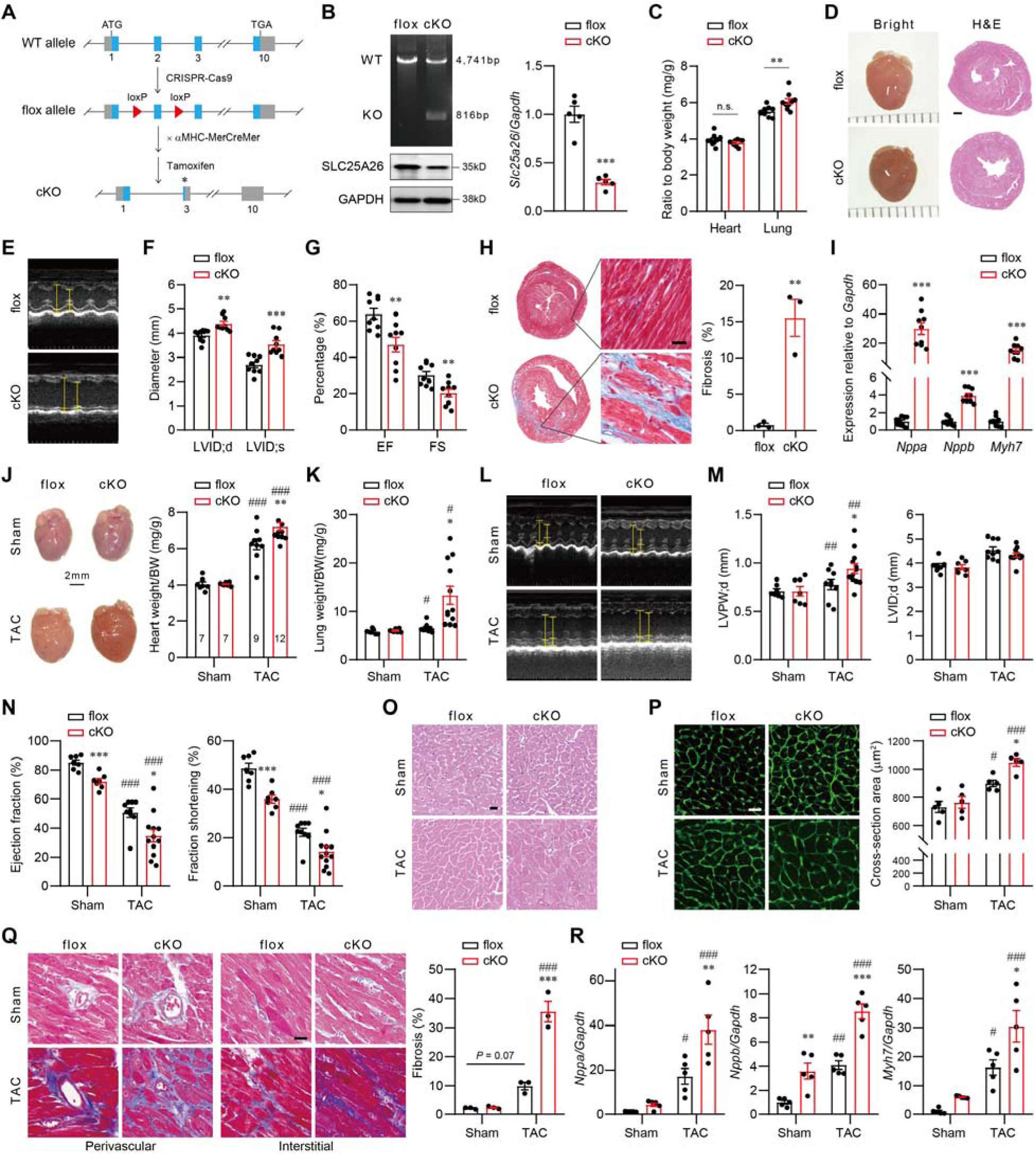
SLC25A26 is required for normal heart function and its deficiency exacerbates TAC-induced cardiac hypertrophy. **A**, Schematic illustrating the generation of *Slc25a26* cardiac-specific knockout (cKO) mouse line. WT, wildtype. **B**, Validation of *Slc25a26* cKO by genotyping (left upper) and expression assessments at protein (left lower) and mRNA (right) levels. *N* = 5. **C**, Effects of *Slc25a26* cKO on heart and lung weights twenty weeks after Tamoxifen administration. *N* = 9. **D**, Representative heart morphology images (left) and low-magnification H&E-stained images (right) of *Slc25a26*-flox and *Slc25a26*-cKO hearts. Scale, 0.5mm. **E** through **G**, Representative Echocardiography images (**E**), quantification analyzed left ventricular end diastolic/systolic internal dimensions (LVID;d/LVID;s; **F**), and quantification analyzed left ventricular ejection fraction (EF) and fraction shortening (FS) (**G**) of *Slc25a26*-flox and *Slc25a26*-cKO hearts twenty weeks after Tamoxifen administration. *N* = 9. **H**, Representative Masson trichrome staining images (left) and quantification data (right) showing myocardial fibrosis in *Slc25a26*-flox and *Slc25a26*-cKO hearts twenty weeks after Tamoxifen administration. Scale, 20μm; *N* = 3. **I**, Effects of *Slc25a26* cKO on the expression of hypertrophic biomarkers natriuretic peptide A (*Nppa*), natriuretic peptide B (*Nppb*), and myosin heavy chain 7 (*Myh7*) measured by qRT-PCR. *N* = 9. **J**, Representative heart morphology images (left) and summarized heart weight/body weight (BW) ratios (right) showing the impact of *Slc25a26* cKO on TAC-induced cardiac hypertrophy. *N* = 7∼12 as labeled on bars. **K**, Impact of *Slc25a26* cKO on the lung weight/BW ratio in sham and TAC mice. *N* = 7∼12. **L** through **N**, Representative Echocardiography images (**L**), quantification analyzed end-diastolic left ventricular posterior wall thickness (LVPW;d) and LVID;d (**M**), and quantification analyzed EF and FS (**N**) showing the impact of *Slc25a26* cKO on TAC-induced cardiac morphology and function. *N* = 7∼12 as mentioned above. **O**, Representative H&E-stained images of *Slc25a26*-flox and *Slc25a26*-cKO hearts with sham or TAC surgeries. Scale, 20μm. **P**, Representative Wheat germ agglutinin (WGA) staining images (left) and quantification data (right) showing the cross-sectional cardiomyocyte area of *Slc25a26*-flox and *Slc25a26*-cKO hearts with sham or TAC surgeries. Scale, 20μm. *N* = 5. **Q**, Representative Masson trichrome staining images (left) and quantification data (right) showing myocardial fibrosis of *Slc25a26*-flox and *Slc25a26*-cKO hearts with sham or TAC surgeries. Scale, 20μm; *N* = 3. **R**, Effects of *Slc25a26* cKO on TAC-induced expressions of *Nppa*, *Nppb*, and *Myh7*. *N* = 5. Data are mean ± SEM. n.s., not significant, *p < 0.05, **p < 0.01, ***p < 0.001 vs. flox; ^#^*P* < 0.05, ^##^*P* < 0.01, ^###^*P* < 0.001 vs. sham, (**B**, **C**, **F**, **G**, **H** and **I**, unpaired Student’s *t* test; **J**, **K**, **M**, **N**, **P**, **Q** and **R**, two-way ANOVA with Tukey’s multiple comparison test).

### SLC25A26 Suppresses Cardiac Hypertrophy

To evaluate the impact of SLC25A26 on pathological cardiac hypertrophy, we subjected the *Slc25a26*-cKO mice (5w after tamoxifen administration) to TAC surgery. Loss of *Slc25a26* significantly elevated the TAC-induced heart weight gain (Figure 2J), and caused severe pulmonary edema in the TAC group (Figure 2K). Echocardiography detected an increased left ventricular posterior wall (LVPW) thickness after TAC, without any change in LVID, in the *Slc25a26*-cKO hearts compared with the *Slc25a26*-flox hearts (Figures 2L and 2M). Moreover, deletion of *Slc25a26* exacerbated the TAC-induced contractility impairment and accelerated the development of heart failure (Figure 2N). At the histological level, *Slc25a26* cKO aggravated the TAC-induced cardiomyocyte enlargement (Figures 2O and 2P) and myocardial fibrosis (Figure 2Q). At the molecular level, the post-TAC inductions of *Nppa*, *Nppb*, and *Myh7* were further potentiated by *Slc25a26* deficiency (Figure 2R). Consistently, *Slc25a26* haploinsufficiency also exhibited worsened hypertrophic pathologies after TAC (Figures S2D through S2J). These data highlight a protective role of SLC25A26 against cardiac hypertrophy.

To examine the inhibitory effect of SLC25A26 on cardiac hypertrophy, we reconstructed adeno-associated virus (AAV9) to overexpress *Slc25a26* in the heart two weeks before TAC surgery (Figures 3A and 3B). *Slc25a26* overexpression significantly restricted the TAC-induced heart weight gain compared with the AAV9-*Vector* control group (Figure 3C). Echocardiography showed that the post-TAC enlargements of LVPW thickness and LVID were significantly blunted by *Slc25a26* overexpression (Figures 3D and 3E); and the post-TAC declines of EF and FS were significantly rescued by *Slc25a26* overexpression (Figure 3F). Moreover, *Slc25a26* overexpression significantly attenuated the cardiomyocyte enlargement (Figure 3G), myocardial fibrosis (Figure 3H), and the induction of pathological genes after TAC (Figure 3I).

**Figure 3.**
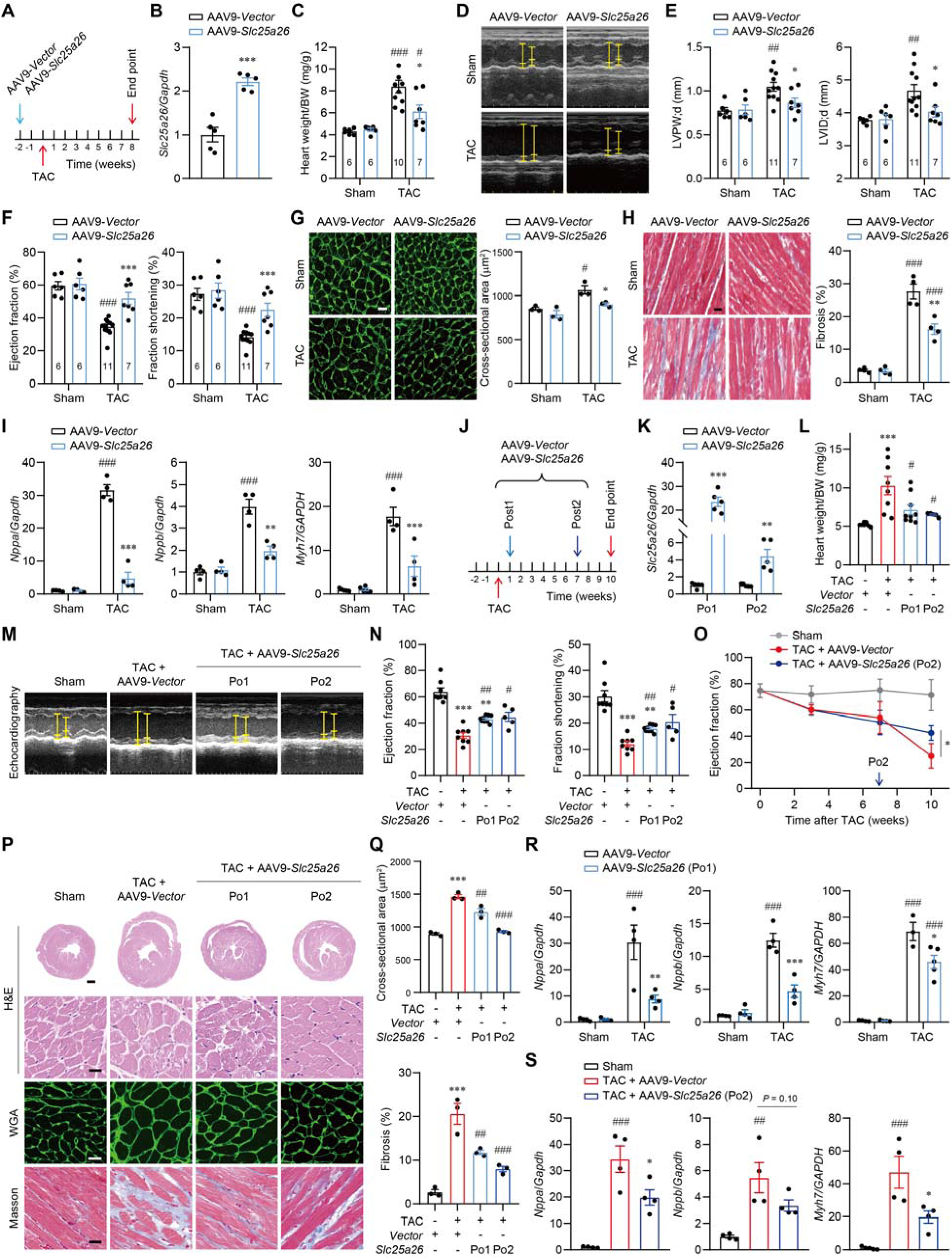
*Slc25a26* overexpression mitigates pathological cardiac hypertrophy. **A**, Injection schedule of reconstructed adeno-associated virus serotype 9 (AAV9) to overexpress *Slc25a26* in the heart before TAC. **B**, Validation of *Slc25a26* overexpression using qRT-PCR. *N* = 5. **C**, Effects of *Slc25a26* overexpression on TAC-induced heart weight gain. *N* = 6∼10 as labeled on bars. **D** through **F**, Representative Echocardiography images (**D**), quantification analyzed LVPW;d and LVID;d (**E**), and quantification analyzed EF and FS (**F**) showing the impact of pre-TAC *Slc25a26* overexpression on TAC-induced cardiac morphological and functional changes. *N* = 6∼10 as labeled on the bars. **G**, Representative WGA staining images (left) and quantification data (right) showing the impact of *Slc25a26* overexpression on TAC-induced cell size enlargement. Scale, 20μm. *N* = 3. **H**, Representative Masson trichrome staining images (left) and quantification data (right) showing the impact of *Slc25a26* overexpression on TAC-induced myocardial fibrosis. Scale, 20μm. *N* = 4. **I**, Impact of *Slc25a26* overexpression on TAC-induced expression of cardiac hypertrophy biomarkers measured by qRT-PCR. *N* = 5. **J**, AAV9 injection schedules to overexpression *Slc25a26* after TAC in two strategies: Post1/Po1 (1w post-TAC) and Post2/Po2 (7w post-TAC). **K**, Validation of *Slc25a26* overexpression in two post-TAC strategies using qRT-PCR. *N* = 5. **L**, Effects of *Slc25a26* overexpression at two post-TAC schedules on TAC-induced heart weight gain. *N* = 5∼9 as indicated by dots. **M** and **N**, Representative Echocardiography images (**M**) and quantification analyzed EF and FS (**N**) showing the impacts of post-TAC *Slc25a26* overexpression on TAC-induced cardiac dysfunction. *N* = 5∼9 as indicated by dots. **O**, Time-course impact of *Slc25a26* overexpression on TAC-induced EF decline in the Po2 strategy. *N* = 5∼8 as mentioned above. **P**, Representative H&E, WGA and Masson staining images of sham and TAC hearts with or without *Slc25a26* overexpression in Po1 and Po2 strategies. Scale, 20μm. **Q**, Quantification data of cross-sectional cardiomyocyte area (upper) and myocardial fibrosis (bottom). *N* = 3. **R** and **S**, Effects of *Slc25a26* overexpression on TAC-induced expressions of *Nppa*, *Nppb* and *Myh7* in Po1 (**R**) and Po2 (**S**) strategies. *N* = 4. Data are mean ± SEM. *p < 0.05, **p < 0.01, ***p < 0.001 vs. AAV9-*vector* or sham; ^#^*P* < 0.05, ^##^*P* < 0.01, ^###^*P* < 0.001 vs. sham or TAC+AAV9-*vector*, (**B** and **K**, unpaired Student’s t test; **L**, **N**, **O**, **Q** and **S**, one-way ANOVA with Tukey’s multiple comparison test; **E**, **F**, **G**, **H**, **I** and **R**, two-way ANOVA with Tukey’s multiple comparison test).

We then challenged whether SLC25A26 would be sufficient to deliver therapeutic benefits by overexpressing *Slc25a26* after TAC in two injection schedules: 1w post-TAC and 7w post-TAC (Figures 3J and 3K). In both strategies, *Slc25a26* overexpression significantly suppressed the TAC-induced heart weight gain (Figure 3L) and restored the declines of EF and FS (Figures 3M and 3N). Notably, *Slc25a26* overexpression at 7w post-TAC significantly rescued the functional decline of the failing hearts (Figure 3O). Moreover, the TAC-induced cardiomyocyte enlargement (Figure 3P), myocardial fibrosis (Figure 3Q), and the induction of pathological markers (Figures 3R and 3S) were all substantially reversed by post-TAC *Slc25a26* overexpression. These data suggest a promising SLC25A26-based gene therapy to treat cardiac hypertrophy.

In line with the *in vivo* findings, the PE-induced hypertrophy of NRVMs was significantly exacerbated by silencing *Slc25a26*, but alleviated by its overexpression (Figures S3A through S3F).

### SLC25A26 Negatively Regulates Translation Activity

To explore the underlying mechanisms, we performed a transcriptome analysis. Principal component analysis showed that *Slc25a26* deficiency caused a generally pro-hypertrophic transcriptome reprogramming (Figure 4A). Gene-concept network analysis highlighted the pathways of aminoacyl-tRNA ligase activity and tRNA aminoacylation for protein translation among the top enriched functional terms affected by *Slc25a26* deletion (Figure 4B). In detail, nearly all cytosol aminoacyl-tRNA synthetases, but not their mitochondrial counterparts, were substantially upregulated in the *Slc25a26*-cKO hearts compared with the *Slc25a26*-flox controls (Figure 4C). Concurrently, gene set enrichment analysis revealed a remarkable upregulation of ribosome-biogenesis-associated genes (Figure 4D), suggesting a crucial role of SLC25A26 in translational regulation.

**Figure 4.**
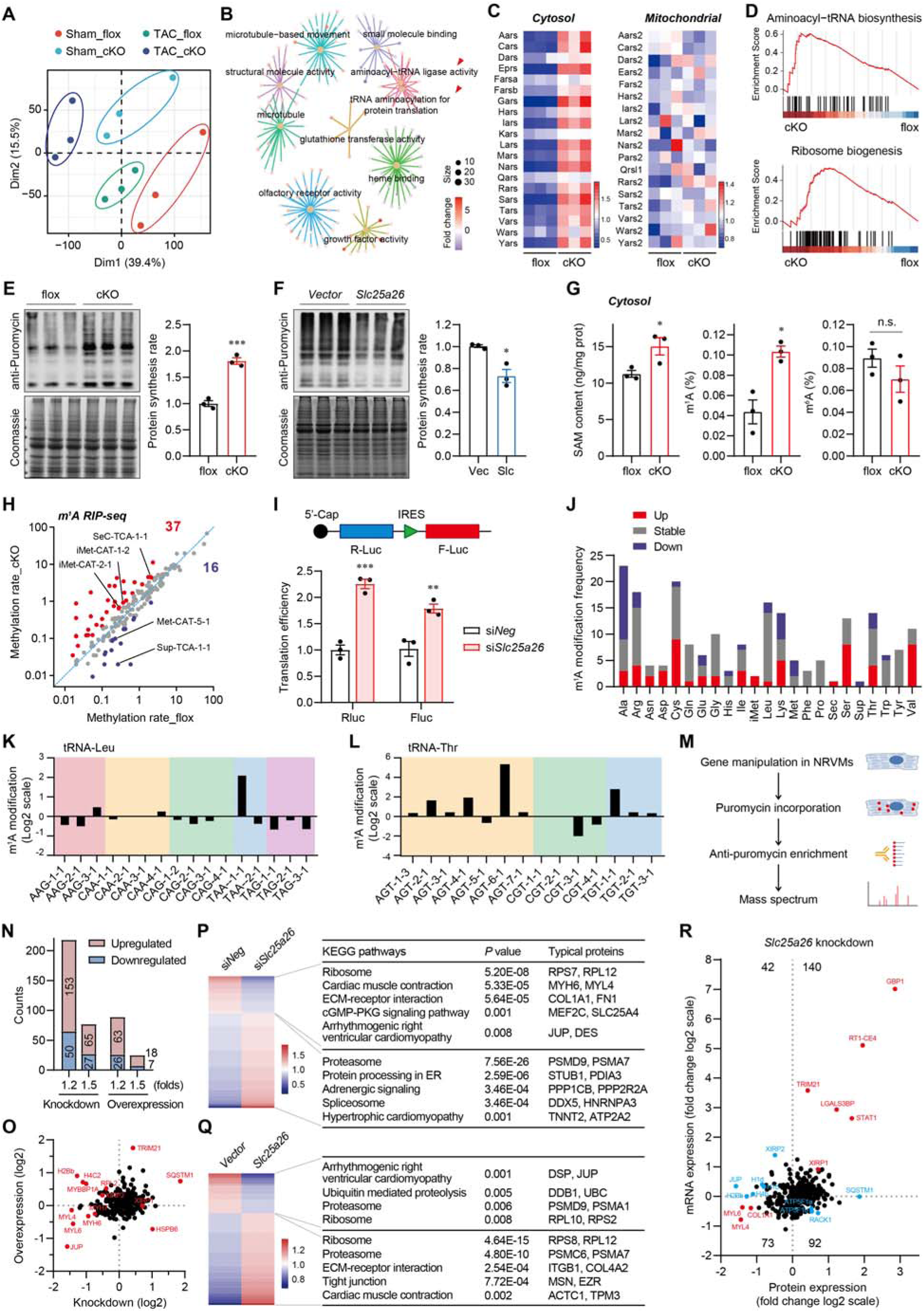
SLC25A26 regulates translation via SAM-mediated tRNA m^1^A modification. **A**, Principal component analysis (PCA) of the RNA-seq data from *Slc25a26*-flox (flox) and *Slc25a26*-cKO (cKO) hearts with sham or TAC surgeries. **B**, Gene concept analysis showing the enriched functional items in the gene profile comparison between flox and cKO hearts. **C**, Heatmap showing the expressions of cytosol (left) and mitochondrial (right) tRNA synthetases in flox and cKO hearts. **D**, Gene set enrichment analysis (GSEA) showing the upregulation of genes related to aminoacyl-tRNA biosynthesis and ribosome biogenesis after *Slc25a26* deletion. **E** and **F**, Puromycin incorporation immunoblots (left) and quantification data (right) showing the protein synthesis rate (normalized to Coomassie blue staining) in the hearts with *Slc25a26* cKO (**E**) or overexpression (**F**). Slc, AAV9-*Slc25a26*; Vec, AAV9-vector. *N* = 3. **G**, ELISA measurements for SAM (left), m^1^A (middle) and m^6^A (right) in the cytoplasmic fraction of flox and cKO hearts. *N* = 3. **H**, Plot of methylated tRNAs detected by m^1^A RNA immunoprecipitation sequencing (RIP-seq) in flox and cKO hearts. **I**, Diagram of the dual luciferase reporter gene plasmid structure (upper) and the impact of *Slc25a26* knockdown on translation initiation efficiency from 5’ cap or internal ribosome entry site (IRES) (lower). *N* = 3. J, m^1^A modification frequency aligned to tRNA variants. **K** and **L**, The m^1^A modification level of tRNA^Leu^ (**K**) and tRNA^Thr^ (**L**) variants detected by m^1^A RIP-seq. **M**, Flowchart of the nascent translatome profiling using a mass-spectrum-based technique for the puromycin-incorporated peptides enriched by immunoprecipitation. **N**, Protein counts detected by immunoprecipitation-mass-spectrum using anti-Puromycin antibody in PE-treated NRVMs with *Slc25a26* knockdown or overexpression. **O**, Plot of puromycin-incorporated proteins from PE-treated NRVMs with *Slc25a26* knockdown or overexpression. **P** and **Q**, Heatmap (left) and gene ontology (GO; right) analyses showing the functional clusters of altered proteins by *Slc25a26* knockdown (**P**) or overexpression (**Q**). **R**, Plot of the changes of translating proteins affected by *Slc25a26* knockdown in comparison with their corresponding mRNA alterations from the RNA-seq analysis. Data are mean ± SEM. n.s., not significant, *p < 0.05, **p < 0.01, ***p < 0.001 vs. flox or si*Neg*, (**E**, **F**, **G** and **I**, unpaired Student’s t test).

We then performed an *in vivo* puromycin incorporation assay to directly assess translation activity. The heart presented the lowest protein synthesis rate among the organs tested (Figure S4A). The protein synthesis rate was significantly accelerated in the *Slc25a26*-cKO hearts but suppressed in the hearts with *Slc25a26* overexpression, compared with corresponding controls (Figures 4E and 4F). Consistently, silencing *Slc25a26* in NRVMs significantly accelerated the protein synthesis rate and further enhanced its activation by PE (Figure S4B), while *Slc25a26* overexpression exhibited opposite effects (Figure S4C). However, *Slc25a26* knockdown failed to re-activate translation when the SAM biosynthesis was inhibited by AG-270, which displayed robust suppression on basal translation activity (Figures S4D and S4E). On the other hand, SAM supplementation largely rescued the inhibitory effect of *Slc25a26* overexpression on translation (Figure S4F). These data suggest that the translation-repressing role of SLC25A26 relies on its function as a SAM transporter.

### SLC25A26 Modulates tRNA m^1^A Profiles and Translation Initiation

Since SAM-dependent tRNA methylation is critical for its maturation and the decoding process during translation,^47, 48^ we hypothesized that SLC25A26-mediated cytoSAM containment might regulate translation through modulating tRNA methylation. In the *Slc25a26*-cKO hearts, an increase in cytoSAM did not affect the N(6)-methyladenosine (m^6^A) level of total RNAs, but significantly enhanced the level of m^1^A (Figure 4G), a modification that mainly occurs on tRNAs.^47–49^ Then we performed RNA immunoprecipitation sequencing (RIP-seq) using an m^1^A-specific antibody to profile the tRNA m^1^A landscape at single-nucleotide resolution. Out of the 400 predicted mouse tRNA isotypes, we identified 202 tRNAs with detectable m^1^A modification (Table S1), among which 37 tRNAs were found to be enhanced in m^1^A modification in the *Slc25a26*-cKO hearts compared with the *Slc25a26*-flox controls, versus only 16 tRNAs were downregulated (Figure 4H). Interestingly, *Slc25a26* deficiency specifically enhanced the m^1^A modification on tRNA^iMet^, but not on its methionine elongation counterpart (tRNA^Met^; Figure 4H), suggesting that the methylation sensitivity of tRNA^iMet^ to SLC25A26-mediated cytoSAM containment governs translation initiation. Indeed, silencing *Slc25a26* in NRVMs significantly enhanced translation initiation efficiency from both 5’-cap and internal ribosome entry sites (IRES) in a luciferase reporter system (Figure 4I).

### SAM-sensitive tRNA m^1^A Patterns Remodel Cardiac Translatome

Status of the tRNA pool determines codon usage that influences the translating rate of individual mRNAs.^32, 50^ We found that the m^1^A modifications after *Slc25a26* deletion were diversified among different tRNA categories (Figure 4J), as well as among different tRNA isotypes for certain amino acids (Figures 4K and 4L). For instance, the m^1^A levels were largely elevated in corresponding tRNAs of Asn, Asp, Cys, Ser, Val and Sec (selenocysteine), but were reduced in those for Ala, His, Trp and Sup (stop codon) after *Slc25a26* knockout (Figure 4J).

To characterize the impact of the cytoSAM-sensitive tRNA m^1^A modifications on translation dynamics, we performed a label-free proteomics analysis for the enriched puromycin-incorporated nascent peptides from NRVMs with different *Slc25a26* manipulations (Figure 4M). We identified 4262 peptides corresponding to 713 proteins, among which 203 proteins were altered by *Slc25a26* knockdown and 92 proteins were altered by its overexpression (Figure 4N). Surprisingly, heart-specific proteins, such as myosin heavy chain (MYH) 6/7, myosin light chain (MYL) 4/6, and Xin actin binding repeat containing (XIRP) 2, were enriched in the downregulated proteins after *Slc25a26* knockdown (Figure 4O). Whereas proteins related to ribosome and cardiac muscle contraction were suppressed by *Slc25a26* knockdown, proteasome-related proteins were highly enriched in the upregulated proteins (Figure 4P). Slc25a26 overexpression generally showed an opposite regulation of these pathways (Figure 4Q). These data suggest a compensatory response to reduce ribosome biogenesis and to augment protein degradation upon *Slc25a26* deficiency. When mapped to the transcriptional changes, roughly 61% proteins, such as XIRP1, MYL4, MYL6, guanylate binding protein 1, signal transducer and activator of transcription 1, tripartite motif containing 21, and collagen type I alpha 1, showed consistent alterations with their mRNA levels; whereas 39% proteins, such as XIRP2, sequestosome 1 (SQSTM1), receptor for activated C kinase 1, and junction plakoglobin, could not be interpreted by their transcriptional alterations (Figure 4R). These data suggest that SLC25A26-mediated biased codon usage remodels cardiac translatome.

### tRNA m^1^A Modification Contributes to SLC25A26-mediated Translational Control

When aligning all the m^1^A sites together, we found that the SAM-sensitive m^1^A modification mainly occurred within the T-arm and the acceptor region of tRNAs, particularly at A58 and A75 (the last nucleotide for esterification with cognate amino acids) (Figure 5A). Typical m^1^A-modified sites were illustrated in tRNA^iMet^, tRNA^Gln^, and tRNA^Thr^ (Figure 5B). However, two tRNA categories, tRNA^Glu^ and tRNA^Gly^, exhibited distinct m^1^A modification patterns specifically in the D arm (Figure 5C). The m^1^A75 modification has not been reported before, but the m^1^A58 modification is well-known for its critical role in tRNA maturation and coding.^47–49^ TRMT6 and TRMT61A function as the methyltransferases for A58 of tRNAs;^51–53^ however, only TRMT61A could be verified necessary for maintaining the normal translation activity in NRVMs (Figures 5D and 5E). Silencing *Trmt61a* significantly reversed the acceleration of the protein synthesis rate by *Slc25a26* knockdown (Figure 5F). Moreover, AAV9-shRNA-mediated *in vivo* knockdown of *Trmt61a* significantly blunted the accelerated protein synthesis rate in the *Slc25a26*-cKO hearts (Figures S5A and S5B), albeit that it did not rescue the TAC-induced heart failure (Figures S5C and S5D). On the other hand, m^1^A58 can be demethylated by *AlkB* homolog 1 (ALKBH1) and ALKBH3.^54–56^ Whereas silencing each gene could both enhance translation activity (Figures 5G and 5H), silencing *Alkbh3* displayed more robust recovery of the restricted translation after *Slc25a26* overexpression than silencing *Alkbh1* (Figures 5I and 5J). These data suggest that tRNA m^1^A modification mediates the translational regulation by SLC25A26.

**Figure 5.**
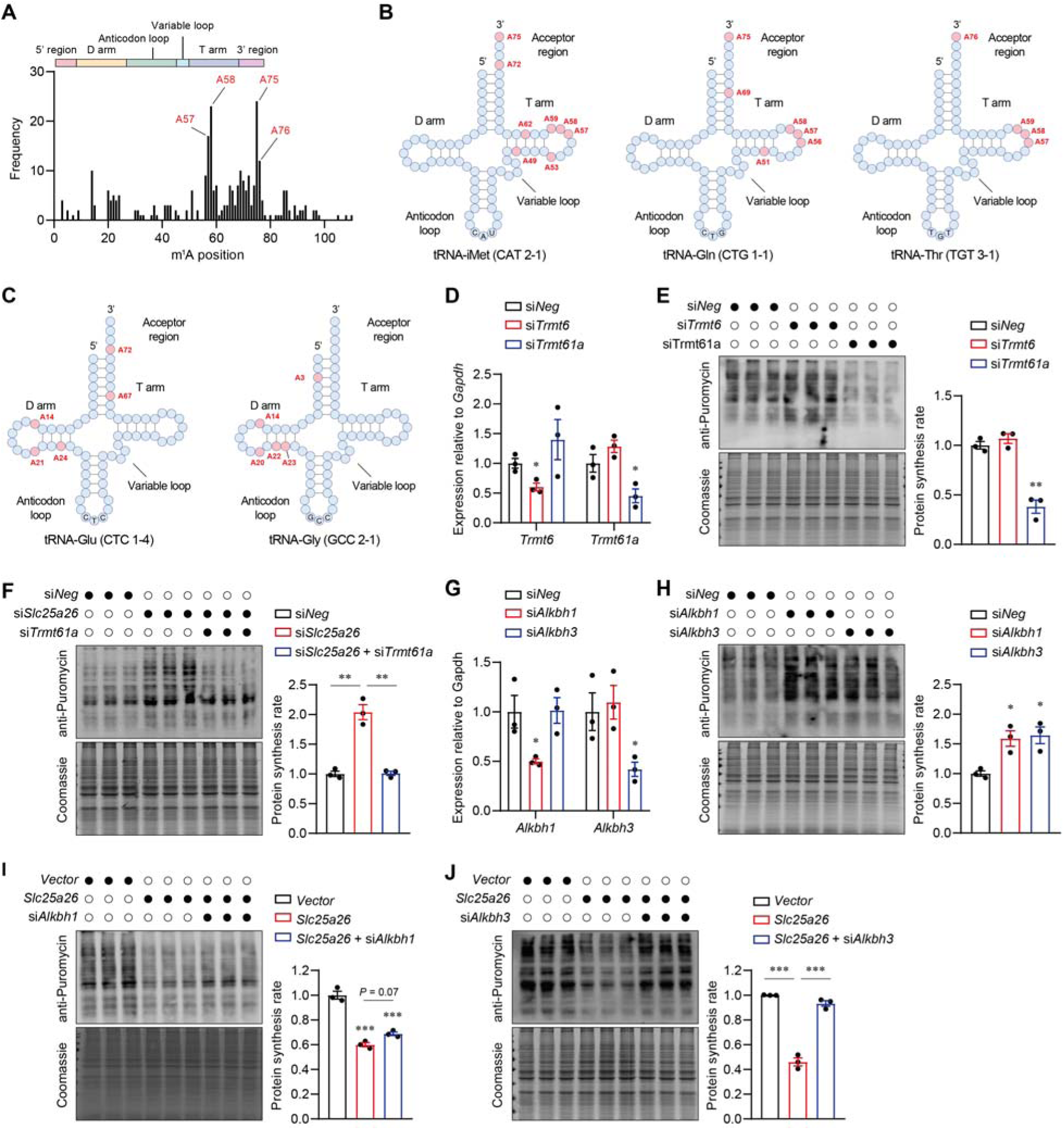
tRNA m^1^A58 modification mediates the translational control by SLC25A26. **A**, Frequency of m^1^A modification in aligned tRNAs at single-nucleotide resolution. **B**, Schematic illustrating the typical m^1^A-modified adenosines in the tRNA^iMet^, tRNA^Gln^ and tRNA^Thr^. **C**, Schematic illustrating the m^1^A modification patterns in thetRNA^Glu^ and tRNA^Gly^. **D**, Validation of the knockdown efficiency of siRNAs targeting tRNA methyltransferases TRMT6 or TRMT61A in NRVMs using qRT-PCR. *N* = 3. **E**, Puromycin incorporation immunoblots (left) and quantification data (right) showing the impact of *Trmt6* or *Trmt61a* knockdown on protein synthesis rate in NRVMs. *N* = 3. **F**, Puromycin incorporation immunoblot (left) and quantification data (right) showing the impact of *Trmt61a* knockdown on protein synthesis rate in NRVMs with *Slc25a26* knockdown. *N* = 3. **G**, Validation of the knockdown efficiency of siRNAs targeting m^1^A demethylases ALKBH1 or ALKBH3 in NRVMs using qRT-PCR. *N* = 3. **H**, Puromycin incorporation immunoblots (left) and quantification data (right) showing the impact of *Alkbh1* or *Alkbh3* knockdown on protein synthesis rate in NRVMs. *N* = 3. **I** and **J**, Puromycin incorporation immunoblots (left) and quantification data (right) showing the impacts of *Alkbh1* knockdown (**I**) or *Alkbh3* knockdown (**J**) on protein synthesis rate in NRVMs with adenovirus-mediated *Slc25a26* overexpression. *N* = 3. Data are mean ± SEM. *p < 0.05, **p < 0.01, ***p < 0.001, (**D**, **E**, **F**, **G**, **H**, **I** and **J**, one-way ANOVA with Tukey’s multiple comparison test).

### mitoSAM Maintains Mitochondrial Fitness for Optimal Energy Production

SAM-dependent methylation reactions are crucial for the normal function of mitochondria,^57^ so we became curious about the impact of SLC25A26 on mitochondrial biology. The mitoSAM level in the heart was reduced by *Slc25a26* cKO but increased by its overexpression (Figure 6A). Nuclear DNA represents a primary methylation substrate sensitive to SAM abundance;^43^ however, it remains under debate whether the mitochondrial DNA (mtDNA) can be methylated.^58^ To address this question, we performed mass spectrometry with purified mtDNA, and found that SLC25A26-mediated mitoSAM accumulation significantly increased the level of 5-methyl-2’-deoxycytidine (5-mdC) with marginal effects on 5-(hydroxymethyl)-2’-deoxycytidine (5-hmdC) or N6-methyl-2’-deoxyadenosine (6-mdA) in mtDNA (Figure 6B). ELISA analysis also confirmed that the 5-mdC amount of mtDNA was reduced in the *Slc25a26*-cKO hearts but augmented in the hearts with *Slc25a26* overexpression, compared with corresponding controls (Figure 6C). Moreover, *Slc25a26* overexpression in NRVMs increased the 5-mdC level in mtDNA but reduced that in the nuclear DNA (Figure S6A). Nevertheless, *Slc25a26* manipulations did not influence the mitochondria copy number (Figures 6D and S6B).

**Figure 6.**
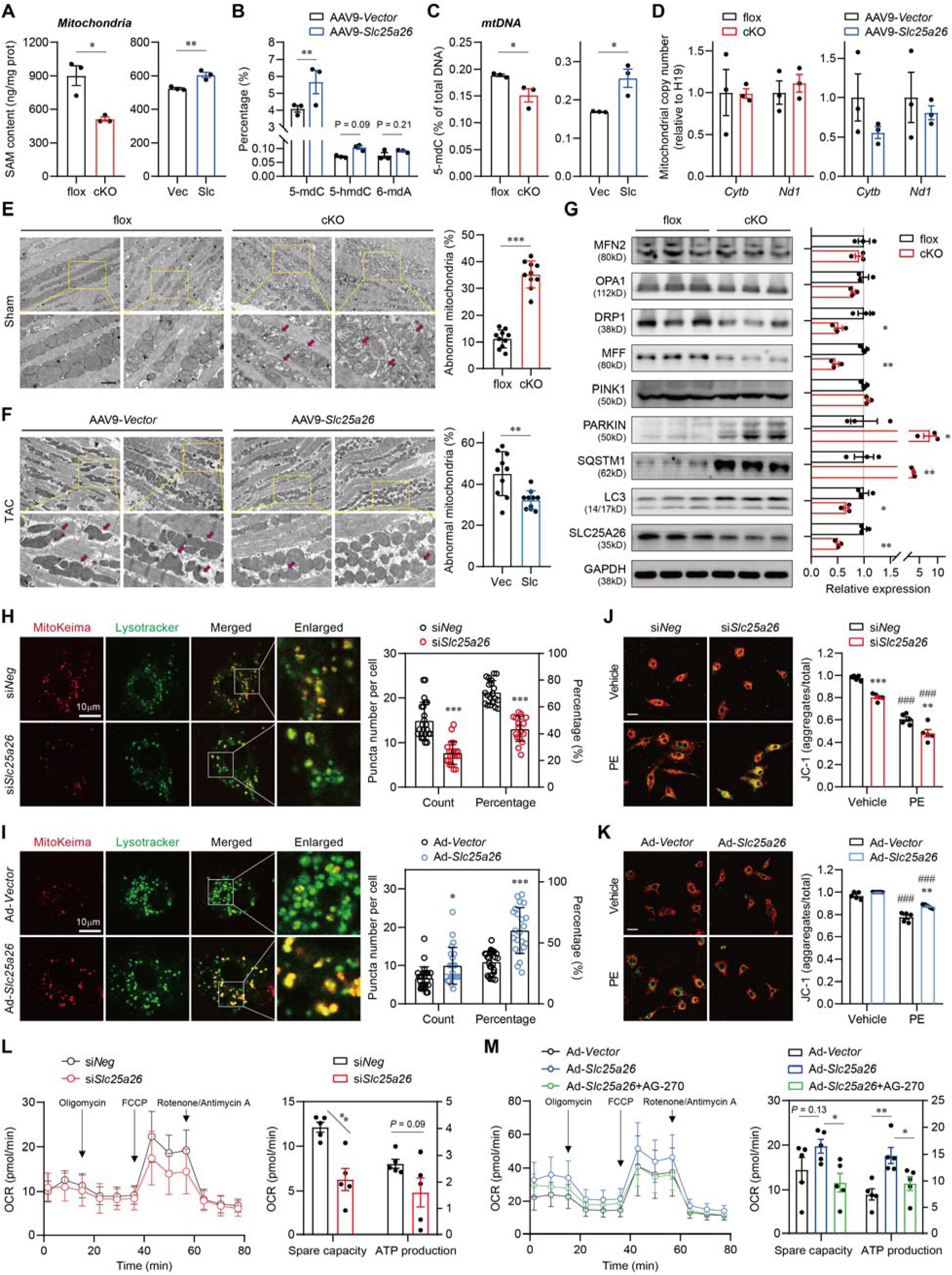
SLC25A26/mitoSAM maintains mitochondrial fitness for optimal energy production. **A**, SAM contents in isolated mitochondria from the hearts with *Slc25a26* cKO (left) or overexpression (right) measured by ELISA. *N* = 3. **B**, Impact of *Slc25a26* overexpression on the methylation of mitochondrial deoxyribonucleotides, including 5-methyl-2’-deoxycytidine (5-mdC), 5-(hydroxymethyl)-2’-deoxycytidine (5-hmdC) and N6-methyl-2’-deoxyadenosine (6-mdA), measured by liquid chromatography-mass spectrometry (LC-MS). *N* = 3. **C**, ELISA measurement for mtDNA 5-mdC from the hearts with *Slc25a26* cKO (left) or *Slc25a26* overexpression (right). *N* = 3. **D**, mtDNA copy number calculated by the expressions of mitochondrial *Cytb* and *Nd1* in contrast to the nuclear *H19* using qRT-PCR. *N* = 3. **E** and **F**, Transmission electronic microscopy showing mitochondrial morphology (left) and quantified abnormal mitochondria percentage (right) in the hearts with *Slc25a26* cKO (sham; **E**) or *Slc25a26* overexpression (post-TAC; **F**). *N* = 3. **G**, Immunoblots (left) and quantification data (right) of proteins involved in mitochondrial dynamics in flox and cKO hearts. *N* = 3. **H** and **I**, Representative fluorescence images (left) and quantification (right) of MitoKeima and Lysotracker showing mitophagy in NRVMs with *Slc25a26* knockdown (**H**) or *Slc25a26* overexpression (**I**). Scale, 10μm; *N* = 3. **J** and **K**, Representative JC-1 staining images (left) and quantification (right) showing mitochondrial membrane potential in NRVMs with *Slc25a26* knockdown (**J**) or overexpression (**K**). Scale, 20μm; *N* = 5. **L** and **M**, Oxygen consumption rate (OCR) of NRVMs with *Slc25a26* knockdown (**L**) or overexpression (**M**) measured by Seahorse. AG-270, 3μM. *N* = 5. Data are mean ± SEM. *p < 0.05, **p < 0.01, ***p < 0.001 vs. flox or AAV9-*vector* or si*Neg* or Ad-*vector*; ^#^*P* < 0.05, ^##^*P* < 0.01, ^###^*P* < 0.001 vs. vehicle, (**A**, **B**, **C**, **D**, **E**, **F**, **G**, **H**, **I** and **L**, unpaired Student’s t test; M, one-way ANOVA with Tukey’s multiple comparison test; **J** and **K**, two-way ANOVA with Tukey’s multiple comparison test).

Transmission electron microscope showed that *Slc25a26* deficiency significantly impaired the mitochondrial morphology in the heart (Figure 6E), whereas *Slc25a26* overexpression significantly ameliorated the severity of mitochondrial damage after TAC (Figure 6F). To explore the underlying mechanism, we screened the expression of proteins related to mitochondrial dynamics. Whereas the fusion-related proteins mitofusin 2 (MFN2) and mitochondrial dynamin-like GTPase (OPA1) did not change after *Slc25a26* deletion, the fission-related proteins dynamin-related protein 1 (DRP1) and mitochondrial fission factor (MFF) were significantly down-regulated (Figure 6G). Moreover, *Slc25a26* deficiency caused significant accumulation of mitophagy-related proteins Parkin and SQSTM1 (also known as p62), but inhibited microtubule associated protein 1 light chain 3 (LC3) lipidation (Figure 6G), suggesting a stagnancy of mitophagy. Similar alteration patterns were observed in NRVMs with *Slc25a26* knockdown (Figure S6C). Furthermore, inhibiting SAM biosynthesis by AG-270 caused a significant decrease of MFF and accumulation of SQSTM1 in despite of a complete depletion of Parkin (Figure S6D). Fluorescent imaging with mito-Keima, a mitophagy indicator,^59^ showed that *Slc25a26* knockdown in NRVMs significantly reduced the engulfment of mitochondria by lysosome (Figure 6H), and this process was oppositely promoted by *Slc25a26* overexpression (Figure 6I). These data suggest that the mitochondrial uptake of SAM is required for the implementation of mitochondrial fission and mitophagy, which are sequentially dependent processes for mitochondrial quality control.^60^

To validate the functional outcome, we performed the JC-1 staining to measure the mitochondrial membrane potential. PE-induced decline in mitochondrial membrane potential was significantly exacerbated by *Slc25a26* knockdown (Figure 6J), but reversed by its overexpression (Figure 6K). Consistently, mitochondrial bioenergetics assay (Seahorse) showed that *Slc25a26* knockdown substantially impaired the mitochondrial respiratory capacity (Figure 6L), whereas its overexpression caused an improvement in oxygen consumption rate and ATP production in an AG-270-sensitive manner (Figure 6M). These results suggest that SLC25A26-mediated mitochondrial fitness benefits cardiac bioenergetics, thereby synchronizing the energy production efficacy with the translation status.

## DISCUSSION

Coordination between translation and bioenergetics is prerequisite for cell flexibility in varying environments; however, such a molecular link is still missing.^4–6, 8^ Our study demonstrates an essential role of SLC25A26-mediated SAM compartmentalization in coordinating translational control and mitochondrial quality control in cardiomyocytes, thus providing the first proof to address this long-standing question.

As the principal methyl donor, the fundamental role of SAM in life activities is self-evident. Firstly, the methyl group is the simplest organic structural unit. Secondly, SAM is derived from methionine, the start amino acid for translation initiation. Thirdly, SAM metabolism pathways are highly conserved among organisms.^61^ Nonetheless, our current understanding about the regulation of SAM biology under pathophysiological conditions remains limited. Here we reveal an increased SAM biosynthesis during the development of cardiac hypertrophy, which drives the mitochondrial translocation of SLC25A26 to shuttle excessive cytoSAM into mitochondria. This regulatory circuit manifests a rare feedforward mechanism that provides a reasonable explanation for the relatively low-level cytoSAM compared to mitoSAM in the heart. It is astonishing that endogenous SLC25A26 is not strictly a mitochondrial protein as originally proposed in Chinese hamster ovary (CHO) cells^40^. In cardiomyocytes, the mitochondrial location of SLC25A26 is adjustable depending on the cellular SAM abundance (Figures 1K and 1L). This is supported by a previous report by Wang et al.^62^ that SLC25A26 was partially co-localized with mitochondria and could form hexamer, a possible existence form in cytosol, as analyzed by size-exclusion chromatography.

The fact that the expression of SLC25A26 is mostly enriched in the heart indicates a special need of the cardiomyocyte to balance the levels of cytoSAM and mitoSAM. In fact, the protein synthesis rate in the heart is among the slowest compared with other tissues (Figure S4A). This concept is further supported by previous observations that the turnover of cardiac sarcomere proteins was relatively slow to maintain normal contractile function^10, 20, 63^. Our data showed that the protein synthesis rate was highly sensitive to the expression alteration of SLC25A26 (Figures 4E, 4F, and S4B-S4F), suggesting that the strict translational control in cardiomyocytes might be attributable to the low-level cytoSAM maintained by SLC25A26.

Our findings verify tRNA m^1^A modification to be the key mechanism underpinning the translational control by SLC25A26 and cytoSAM. The m^1^A58 modification is a conserved feature of all tRNA variants, and has been thought to aid tRNA maturation, stability and decoding.^47–49^ Evidence in this study implicates that m^1^A58 is not strictly required for the housekeeping function of the tRNA but can be regulated by SAM availability, and serves as a gearbox to finetune the translation efficiency. The m^1^A58 modification on tRNA^iMet^ has been reported to promote translation initiation, but its pathophysiological relevance in human diseases has been lacking until very recently.^54^ The alteration in m^1^A58 level provides a mechanistic explanation for how proteins are excessively synthesized during cardiac hypertrophy as a result of enhanced SAM biosynthesis. Our data also validated the bidirectional regulation of m^1^A58 by the writer TRMT61A and the erasers ALKBH1/3.^52–54^ These modulators provide a comprehensive tool box to modify the translation status by manipulating the level of m^1^A58.

Moreover, we identified m^1^A75 for the first time as a novel methylation event that was remarkably abundant in tRNAs (Figure 5A). Considering that this is the last nucleotide for esterification with cognate amino acids, m^1^A75 might have crucial functions in translational regulation.

Beyond translation initiation, we also provide proof-of-concept evidence for the biased codon usage derived from diversified tRNA m^1^A modifications in mammalian cells. The uneven use of synonymous codons in the transcriptome serves as a secondary genetic code guiding translation efficiency and fidelity.^64^ Synonymous codon usage can have a substantial influence on protein synthesis rates.^65^ Highly expressed genes exhibit improved adaptation of the codons to the tRNA pool.^66^ Li et al.^67^ found that tRNA m^5^C modification stabilizes tRNA^Leu^, contributing to a codon-dependent translation bias to support a pro-cancer translation program. Our data reveal that m^1^A modifications to different tRNA variants and different isotypes of certain tRNA category possess diversified sensitivity to the SAM availability. This mechanism is preferentially leveraged by the cardiomyocytes to tailor the proteome under hypertrophic stress. Nevertheless, how these changes participate in pathogenesis of cardiac hypertrophy requires further validation in details.

Mitochondrial fission enables clearance of dysfunctional mitochondria through mitophagy to maintain the integrity of the energy system.^60^ The high enrichment of SAM makes mitochondria a pool for intracellular SAM, a unique pattern usually adopted by ions (e.g. calcium) rather than metabolites. SLC25A26-mediated increase in mitoSAM is accompanied by enhanced mitochondrial fission, mitophagy and ATP production, suggesting an important role of SLC25A26/mitoSAM in mitochondrial quality control. Although we did not clarify the direct effectors, the regulation of mitochondrial biology by mitoSAM is closely related to its impact on mtDNA methylation. Certain isoforms of DNA methyltransferases can enter mitochondria to catalyze mtDNA methylation, which has been associated with mitochondrial replication, dynamics and bioenergetics.^68, 69^ *Slc25a26* mutations in human cause intra-mitochondrial methylation deficiency, and are associated with neonatal mortality resulting from respiratory insufficiency and hydrops, childhood acute episodes of cardiopulmonary failure, and slowly progressive muscle weakness,^43–45^ suggesting an essential role of SLC25A26-mediated mitochondrial homeostasis during development.

Putting the differential functions of cytoSAM and mitoSAM in the context of cardiac hypertrophy, a rational deduction is that the enhanced SAM biosynthesis and transport are intrinsic protective mechanisms to compensate for the increased contractility and energy demands of cardiomyocytes under pressure overload. However, excessive protein synthesis under long-term stress might exhaust the mitochondria for faster and faster ATP generation, eventually leading to decompensation and the transition from cardiac hypertrophy to heart failure.^70^ In this case, SLC25A26 overexpression is efficient to lower cytoSAM but increase mitoSAM, resulting in translation slowdown and mitochondrial fitness. This scenario provides a promising gene-therapy strategy to kill two birds with one stone.

Taken together, our study demonstrates SLC25A26-mediated SAM compartmentalization as a molecular link between translation control and mitochondrial quality control. Targeting intracellular SAM distribution would be a promising therapeutic strategy to treat cardiac hypertrophy and heart failure.

## Data Availability

RNA-seq data have been deposited in NCBIs Gene Expression Omnibus (GEO) repository (accession: GSE254565 and GSE173737). Online microarray datasets are available from NCBIs GEO repository with corresponding accession codes as described in details in Supplemental Methods. Any additional information reported in this paper is available from the lead contact upon request.

## Nonstandard Abbreviations and Acronyms

4E-BP1: eukaryotic translation initiation factor 4E binding protein 1
5-hmdC: 5-(hydroxymethyl)-2’-deoxycytidine
5-mdC: 5-methyl-2’-deoxycytidine
6-mdA: N6-methyl-2’-deoxyadenosine
AAV9: adeno-associated virus serotype 9
ALKBH1/3: AlkB homolog 1/3
ATP: adenosine triphosphate
cKO: cardiac-specific conditional knockout
cytoSAM: cytoplasmic SAM
DRP1: dynamin-related protein 1
EF: ejection fraction
ELISA: Enzyme-linked immunosorbent assay
FS: fraction shortening
LC3: microtubule associated protein 1 light chain 3
LVID: left ventricular internal diameter
LVPW: left ventricular posterior wall
m1A N1: methylated adenosine
m6A N6: methylated adenosine
MAT2A: methionine adenosyltransferase 2A
MFF: mitochondrial fission factor
MFN2: mitofusin 2
mitoSAM: mitochondrial SAM
mTORC1: mechanistic target of rapamycin kinase complex 1
MYH6/7: myosin heavy chain 6/7
MYL4/6: myosin light chain 4/6
Nppa/Nppb: natriuretic peptide A/B
NRVMs: neonatal rat ventricular myocytes
OPA1: mitochondrial dynamin-like GTPase
RIP-seq: RNA immunoprecipitation sequencing
SAM: S-adenosylmethionine
SLC25A26/SAMC: solute carrier family 25 member 26
SQSTM1: sequestosome 1
TAC: transaortic constriction
TRMT6/61A: tRNA methyltransferase 6/61A
XIRP2: Xin actin binding repeat containing 2

## Acknowledgments

We would like to thank Yingying Guo and Shun Wang from Wuhan University for their assistance on animal experiments. Mito-Keima plasmid was a gift from Dr. Moshi Song’s lab.

## Supplemental Material

Supplemental Methods

Figures S1-S6

Tables S1-S2

Supplemental References

