## Supplemental Methods, Supplemental Tables S1 and S2, Supplemental Figures S1-S6 for "SLC25A26-mediated SAM compartmentalization coordinates translation and bioenergetics during cardiac hypertrophy"

This file contains Supplemental Methods, Figures S1-S6, and Tables S1-S2.

### SUPPLEMENTAL METHODS

#### Human Samples

This study was approved by the Ethics Committee of the Fuwai Hospital Chinese Academy of Medical Sciences, Shenzhen (SP2023133(01)). Human heart samples were collected from two patients with obstructive hypertrophic cardiomyopathy patients during myectomy surgical treatment in Fuwai Hospital Chinese Academy of Medical Sciences, Shenzhen with written informed consents signed in advance.

#### Mice

All experiment procedures with mice were reviewed and approved by the Institutional Animal Care and Use Committee (IACUC) of Renmin Hospital of Wuhan University (No. 20180508) and the IACUC of Fuwai Hospital Chinese Academy of Medical Sciences, Shenzhen (SP2023059), and performed in accordance with the guide for the care and use of laboratory animals published by National Institutes of Health, USA (8^th^ edition). Mice were raised in a specific pathogen free environment (room temperature, 24 ± 3°C; room humidity, 55 ± 5%) with a 12-h light/12-h dark cycle and fed normal chow. *Slc25a26* knockout mice were generated using clustered regularly interspaced short palindromic repeats (CRISPR)-cas9 technology by GemPharmatech Inc. (Nanjing, China). Guide RNAs (gRNAs) flanking the second exon were designed to delete this exon and generate a frame-shift mutation in SLC25A26. The *Slc25a26*^flox/flox^ mouse line was generated by CRISPR-cas9-mediated genome editing to insert two loxP sites flanking the exon 2 of the *Slc25a26* gene (NCBI Reference Sequence NM_026255.5). Then the *Slc25a26*^flox/flox^ mice were bred with the αMHC-MerCreMer mice ^71^ to generate the *Slc25a26*^fl/fl^;αMHC-MerCreMer mice for inducible *Slc25a26* cKO. Deletion of Slc25a26 was realized by injection of Tamoxifen (Sigma-Aldrich, #T5648; 40mg/kg/day; i.p.) for 5 times at the age of 5 weeks old.

#### *In Vivo Slc25a26* Overexpression

For *Slc25a26* overexpression in the heart, we constructed the recombinant adeno-associated virus serotype 9 (AAV9) expressing the Flag-tagged mouse *Slc25a26* under the control of CMV promoter. AAV9-vector was used as a negative control. Male mice with C57BL/6 background were randomly separated into AAV9-vector and AAV9-*Slc25a26* groups and injected with 6 ×10^12^ vector genomes (VG)/kg viruses through the tail vein by three different strategies: 2 weeks before transaortic constriction (TAC), 1 week after TAC, and 7 weeks after TAC. The overexpression of SLC25A26 was validated by quantitative real-time polymerase chain reaction (qRT-PCR) as described below.

#### Transaortic Constriction (TAC) Surgery

TAC surgery was performed as previously described.^72, 73^ Male C57BL/6J mice (8~10 weeks old; weighing 24~26g) were randomly separated into sham and TAC groups, group information was double-blind between the investigator performing the surgery and the data analyst. Mice were anesthetized by intraperitoneal injection with 50mg/kg sodium pentobarbital and supported by a mouse mini-ventilator (Alcott biotech, Shanghai, China). The respiration rate was set to 110breaths/min, tidal volumes to 2ml/min, and inspiration/expiration ratio to 1:2. Body temperature was maintained at 37℃ throughout the experimental procedure using a self-regulating heating pad. After depilation with depilatory cream and routine disinfection, the mouse skin was incised at the junction of the sternum and the second rib. Partial thoracotomy to the second rib was performed under a surgical microscope till clearly seeing the transverse aorta. A 27-gauge needle was placed between the brachiocephalic trunk and left carotid arteries. After aorta ligation with 7-0 silk sutures, the needle was then quickly removed to generate a constriction of 0.4mm in diameter. Then the thoracic cavity was closed layer by layer. In the sham control mice, all procedures were identical except for the ligation of the aorta. Successful constriction of the aorta was validated by ultrasound Doppler in Echocardiography.

#### Echocardiography

Transthoracic echocardiography was performed before and after the TAC surgery using an ultrasound system (Vinno 6, Suzhou, China) as previously described ^73^. Mice were anesthetized with 1.5~2% inhaled isoflurane. M-mode echocardiograms were recorded at the mitral papillary muscle level under the left ventricular short axis views. Left ventricular end systolic/diastolic posterior wall thickness (LVPW;s/LVPW;d), left ventricular end systolic/diastolic inner dimension (LVID;s/LVID;d), left ventricular end systolic/diastolic volume (LVESV/LVEDV), ejection fraction (EF) and fractional shortening (FS) were measured and calculated from at least three consecutive heartbeats according to the American Society of Echocardiography guidelines.

#### Histology

After sacrifice of the mice, the hearts were harvested and dipped in 1M KCl solution to arrest them in diastole status, then rinsed in saline solution and fixed in 4% paraformaldehyde for 24h at room temperature. Subsequently, these hearts were embedded in paraffin using standard histological procedures and sectioned at 5µm. The sections were stained with hematoxylin-eosin (HE) for histopathology, Masson trichrome staining for fibrosis, or wheat germ agglutinin (WGA; Invitrogen, #W11261) for measurement of the cardiomyocyte cross-sectional area. The stained sections were visualized by a confocal microscope (Olympus FV3000, Tokyo, Japan) and analyzed by Image Pro Plus 6.0 software (Media Cybernetics, Bethesda, USA).

#### Electron Microscopy

Heart tissues from different groups of mice were freshly harvested and cut into blocks roughly 1mm^3^. The blocks were fixed with 0.1M Cacodylate buffer (pH7.2) containing 2% paraformaldehyde and 2% glutaraldehyde for 2h at room temperature and then washed twice in 0.1M Cacodylate buffer. The samples were subsequently post-fixed with 1% osmium tetraoxide in 0.1M Cacodylate buffer for 1h at 4°C and then stained with 0.5% uranyl acetate (*p*H 4.0) overnight at 4°C. Samples were then dehydrated by incubation in a graded series of ethanol solutions (30%, 60%, 90% and three times in 100%) prior to be infiltrated with an ethanol/resin mixture (1:1) for 1h at room temperature. Following polymerization overnight at 65°C, 80-nm sections were cut on Leica EM UC7 (Leica Microsystems, Wetzlar, Germany) and picked up on copper grids. The grids were post-stained in uranyl acetate and bismuth subnitrate. The sections were observed in a Hitachi HT7800 (Hitachi, Tokyo, Japan) and micrographs recorded.

#### Cell Culture

Neonatal rat ventricular myocytes (NRVMs) were isolated and cultured as previously described ^74^. Briefly, the hearts were extracted from 1~3d Sprague-Dawley (SD) neonatal rats. After removing the atrium and the large vessels attached to the surface of the heart, ventricles were finely cut into small pieces of about 1mm, and then digested in a solution containing 0.08% type II collagenase (Sigma-Aldrich, #1148090) and 0.125% trypsin (Sigma-Aldrich, #[T2600000](https://www.sigmaaldrich.cn/CN/zh/product/sial/t2600000)) at 37℃ for 3 times (20min each). Cardiomyocytes were separated from fibroblasts by percoll (GE, #17-0891-09) density gradient centrifugation, and cultured in Dulbecco's modified Eagle's medium (DMEM; Hyclone, #SH30243.01) containing 10% (v/v) heat-inactivated fetal bovine serum (FBS; Gibco, #10099141C) and 1% penicillin and streptomycin (Gibco, #15140163). After 24h, the FBS was replaced by 1% insulin–transferrin–selenium (ITS; Cyagen, #10201) for another 24h before further manipulations. Plasmids were transfected into NRVMs by Nanoparticle In Vivo Transfection Reagent (Altogen, #5031) as previously described ^73^, and siRNAs were transfected by Lipofectamine RNAiMAX (ThermoFisher, #13778150). Reagents, including S-adenosylmethionine (SAM; Sigma-Aldrich, #A7007; 1μM), AG-270 (MAT2A inhibitor; MCE, #HY-138630; 3μM), rapamycin (mTOR inhibitor; MCE, #HY-10219; 1μM) were dissolved with dimethylsulfoxide (DMSO) as 1000 times concentrated stock and filtered with 0.22µm membrane for sterilization.

#### Cell size Measurement

Size of NRVMs was measured by WGA staining in 6-well plates. After treatments, NRVMs were washed with PBS and fixed with 4% paraformaldehyde for 15min at room temperature. They were then permeabilized with 0.2% Triton X-100 at room temperature for 10min, and incubated with WGA (5.0µg/mL; Invitrogen, #W11261) for 10min at 37°C, followed by DAPI staining for 10min at room temperature. Images were collected using an inverted fluorescence microscope (Olympus CKX53, Tokyo, Japan) and analyzed by the Image Pro Plus 6.0 software. More than 200 cells in each group were analyzed to calculate the surface area of cardiomyocytes. Representative NRVM images were labeled with α-actinin antibody (1:100, Proteintech, #11313-2-AP) using immunofluorescence as previously described ^73^.

#### Immunofluorescence

Subcellular localization of SLC25A26 in NRVMs was visualized by immunofluorescence for ectopically expressed Flag-tagged SLC25A26. Briefly, NRVMs were cultured on sterile glass slides in 6-well plates. SAM (1μM), PE (50μM) of AG-270 (3μM) were supplemented for 24h after adenoviruses infection for 24h. The cells were then fixed with 4% formaldehyde in PBS for 15min at room temperature, permeabilized with 0.1% Triton X-100 in PBS for 10min, goat serum was blocked at room temperature for 30min, and then incubated with Anti-Flag-488 (1:1000, Abcam) and Anti-Cox1 (1:100, Abcam) at 4°C overnight. The next day, cell samples were incubated with corresponding secondary antibodies for 1h at room temperature, dyed with DAPI for 10min at room temperature, and then washed 5~7 times with PBS before mounting on slides. Images were collected using an inverted fluorescence microscope (Olympus CKX53, Tokyo, Japan) and analyzed by the Image Pro Plus 6.0 software.

#### Mitochondrial Dynamics

NRVMs with *Slc25a26* knockdown, overexpression, or AG-270 (0.3µM) treatment were co-transfected with the mito-Keima plasmid on coverslips in 6-well plates, and co-stained with Lysotracker (Thermo Fisher, #L7526; 100nM) to visualize mitophagy as previously described^75^. The mitochondrial autophagy was measured by counting the number of yellow spots in each cell. The mitochondrial membrane potential was measured in PE-treated NRVMs with *Slc25a26* knockdown or overexpression using an enhanced mitochondrial membrane potential assay kit based on JC-1 (Beyotime, C2003S) according to the manufacturer's protocol. Images were quantified by calculating the aggregates (red) and monomers (green). To measure the mitochondrial copy number, DNA from frozen mouse hearts was extracted and purified using the MagMAX Total Nucleic Acid Isolation Kit (Applied Biosystems, #AM1840) according to the manufacturer's instruction. The quantification of mitochondrial DNA was determined by qRT-PCR using Nd1 and cytochrome b (*Cytb*). Mitochondrial DNA (mtDNA) copy number was expressed relative to nuclear gene H19.

#### Mitochondrial Respiration

NRVMs (5×10^4^) were seeded in the XF96 cell culture plate (Agilent Technologies, CA, USA) and placed in a cell culture incubator at 37 °C, 5% CO_2_ overnight. The culture medium was replaced with XF Base Medium (*p*H 7.4) supplemented with 1.0M Glucose Solution, 100mM Pyruvate Solution, and 200mM Glutamine Solution (Agilent Technologies, CA, USA) after *Slc25a26* knockdown, overexpression, or AG-270 treatment; it was then incubated in a CO_2_-free cell incubator at 37°C for 60min. Oxygen consumption rate（OCR）was measured using the Seahorse XF cell mito stress test kit (Agilent Technologies, CA, USA), according to the manufacturer’s instructions. Oligomycin (1.5μM), FCCP (0.5μM), and rotenone/antimycin A (Rot/AA, 0.5μM) were added to each well; then, the plate was transferred to a Seahorse XF96 analyzer (Agilent Technologies, CA, USA) for analysis. OCR was normalized according to the number of cells.

#### Plasmids

The full-length cDNA sequence of mouse and rat *Slc25a26* (m*Slc25a26* and r*Slc25a26*) were obtained by the National Center for Biotechnology Information (NM_026255.5 and NM_001395605.1, respectively). The pcDNA3.1-mSlc25a26-HA/Flag and pcDNA3.1-rSlc25a26-HA/Flag recombinant plasmids were constructed by cloning the coding region of m*Slc25a26* and r*Slc25a26* into the NheI and XbaI sites of the pcDNA3.1-HA/Flag plasmids. Massive plasmid replication was performed by transforming the plasmids into E.coli DH-5α. m*Slc25a26* was encompassed into the adenovirus or adeno-associated virus vectors to overexpress it in NRVMs or mouse hearts, respectively. Mito-Keima plasmid was a gift from Prof. Moshi Song from Institute of Zoology, Chinese Academy of Sciences.

#### Western Blot

Total proteins were extracted from cells or heart tissues with Radio Immunoprecipitation Assay (RIPA) lysis buffers (Beyotime, #P0013B). Cell lysates and heart homogenates were centrifuged at 12,000×g for 15min. Protein concentration was measured by the BCA method, and equal quantities of protein extracts were loaded on a sodium dodecyl sulfate-polyacrylamide gel electrophoresis (SDS-PAGE), then transferred onto polyvinylidene fluoride (PVDF) membranes (Millipore, #IPVH00010). The membranes were blocked with 5% (w/v) non-fat milk for 1h at room temperature, and incubated with specific primary antibodies overnight at 4°C. The membranes were then incubated with corresponding HRP-linked secondary antibody at room temperature for 1h, and visualized by ECL Western Blotting Detection Kit (ThermoFisher, #32109). Specific primary antibodies included: anti-SLC25A26 (Abclonal, #WS138422), anti-GAPDH (Proteintech, #HRP-60004), anti-Histone H3 (Proteintech, #17168-1-AP), anti-VDAC1 (Abcam, #ab14734), anti-COX1 (Abcam, #ab147005), anti-Puromycin (Sigma-Aldrich, #MABE343), anti-MAT2A (Abcam, #ab154343), anti-MAT2B (Abcam, # ab129176), anti-cTnT (Santa Cruz, #sc-52284), anti-OPA1 (CST, #80471), anti-MFN2 (Abclonal, #WH149521), anti-Drp1 (Abclonal, #A16661), anti-MFF (Cell Signaling Technology, CST; # 84580)，anti-Parkin (CST, #4211)，anti-PINK (CST, #6946)，anti-SQSTM1 (CST, # 8025), anti-LC3 (CST, # 3868).

#### Puromycin Incorporation Assay

Puromycin mimics the aminoacyl-end structure of the amino-acid-tRNA, and can be readily incorporated into nascent polypeptides, thus reflecting a snapshot of ongoing translation events ^73, 76, 77^. For cell experiment, cells were treated with puromycin (1μM) for 30min before harvesting cells, while mice were given 25mg/kg by intraperitoneal injection 3h before being killed, then extracted total proteins, followed by immunoblotting with the anti-Puromycin antibody (Sigma-Aldrich, #MABE343). Protein synthesis rate was calculated by the histograms of Puromycin signal against Coomassie blue staining.

#### Dual-luciferase Assay

Translation initiation was evaluated by the pcDNA3-RLuc-IRES-FLuc system as previously described ^78^. The bicistronic luciferase vectors containing internal ribosome entry site (IRES) elements were generated by subcloning the coding sequence for firefly luciferase (Fluc) from pGEM-LUC (Promega), the coding sequence for renilla luciferase (Rluc) from pRL-CMV (Promega) and IRES sequence of oncogenes into pcDNA3.1 vector. HEK293T cells with or without *Slc25a26* overexpression were transfected with the indicated bicistronic constructs. Dual-luciferase assays were performed after 48h of transfection using the Dual-luciferase reagent (Promega, #E1910) according to the manufacturer’s instructions.

#### RNA Sequencing (RNA-seq) Analysis

Total RNA was extracted from heart tissues with the TRIzol reagent (Invitrogen, #15596018). RNA-seq analysis was performed by the BGISEQ platform of Beijing Genomics Institution (BGI) as previously described ^73, 77^. Three independent biological replicates were sequenced for each group. The different expression genes (DEGs) between groups were calculated using the linear models for microarray data (limma) package in R with a threshold of |Fold change| >1.5 and adjusted P-value < 0.05. Gene ontology (GO) analysis of DEGs was performed by Metascape online database with default parameters. Gene set enrichment analysis (GSEA) was used to screen significantly enriched signalling pathways. Raw data have been uploaded to the Gene Expression Omnibus (GEO, <http://www.ncbi.nlm.nih.gov/geo>) database with the access number GSE254565. To determine the expression of Slc25a26 in TAC-induced cardiac hypertrophy, we also analyzed online microarray datasets, including GSE61177^ref. in 79^, GSE24489^ref. in 80^, GSE37597, GSE6970 ^ref. in^ ^81^, GSE27689^ref. in 82^, GSE18224^ref. in 83^, GSE5500^ref. in 84^, GSE56348^ref. in 85^, GSE50638 and GSE69355^ref. in 86^, downloaded from the GEO database.

#### Label-free Proteomics

To identify the translating proteins regulated by SLC25A26, we enriched the Puromycin-incorporated peptides using anti-Puromycin-based immunoprecipitation and performed the label-free proteomics analysis (PTM Biolabs, Hangzhou, China) using PE-treated NRVMs with *Slc25a26* knockdown or overexpression. Briefly, the pull-down peptides with Puromycin incorporation were reduced with 5mM dithiothreitol for 30min at 56°C and alkylated with 11mM iodoacetamide for 15min at room temperature in darkness. The samples were then diluted by adding 100mM TEAB to urea concentration less than 2M. Trypsin was added at 1:50 trypsin-to-protein mass ratio for the first digestion overnight and 1:100 trypsin-to-protein mass ratio for a second 4h-digestion. Finally, the peptides were desalted by C18 SPE column. The digested peptides were analyzed in Q ExactiveTM HF-X (ThermoFisher) with a nano-electrospray ion source. The resulting MS/MS data were processed using MaxQuant search engine (v.1.6.15.0). Tandem mass spectra were searched against the human SwissProt database (20422 entries) concatenated with reverse decoy database. KEGG analysis was performed by Enrichr online tool (<https://maayanlab.cloud/Enrichr/>).

#### Quantitative Real-time Polymerase Chain Reaction (qRT-PCR)

Total RNA was extracted from cells or heart tissues using TRIzol reagents (Invitrogen, #15596018) and quantified using NanoDrop (ThermoFisher). The cDNAs were synthesized with 1μg RNA using RevertAid First Strand cDNA Synthesis Kit (ThermoFisher, #K1622). qRT-PCR was performed using Ultra SYBR Mixture (Roche, #6924204001) on CFX96M Touch Real-Time PCR Detection System (Roche, Basel, Switzerland). The primer sequences used in this study were listed in Table S2.

#### m^1^A tRNA Immuno-precipitation Sequencing (RIP-Seq)

The m^1^A tRNA RIP-Seq was used to explore the m^1^A modifications in tRNAs as previously described ^87^. The m^1^A RIP-Seq service was provided by CloudSeq Biotech (Shanghai, China). Briefly，total RNA was extracted using Trizol reagent (Invitrogen, CA, USA) following the manufacturer's protocol. Small RNAs (<200nt) were enriched from total RNA using the MirVana Isolation Kit (Thermo Fisher). RNAs without demethylase treatment were used as input. The m^1^A RIP reaction was performed using the m^1^A MeRIP kit from (GenSeq, Inc) according to the manufacturer’s instruction. RNA and m^1^A antibody were co-incubated at 4°C for 2h in IPP buffer [10mM Tris-HCl (*p*H7.5), 150mM NaCl, 0.1% NP-40]. The reaction mixture was further immunoprecipitated with protein A magnetic beads (Thermo Fisher) at 4°C for 2h. Then the bound RNA on the magnetic beads was eluted with free m^1^A adenosine analogue. The samples after IP were divided into two parts, one part was treated with demethylase AlkB [demethylase (+)], and the other part was not treated [demethylase (-)]. Finally, the obtained samples were respectively built into a library using the RNA Builder Kit (NEB) and sequenced on Illumina Nextseq platforms generating short paired-end reads, ranging from 25 to 55bp from each end.

#### Measurement of SAM Contents

Total SAM contents in NRVMs or mouse hearts were measured with an enzyme-linked immunosorbent assay (ELISA) kit (mlbio, China) according to the manufacturer’s instruction. To measure the subcellular SAM contents, NRVMs or heart tissues were fractionated to separate mitochondria, cytosol and nucleus. Samples were homogenized in 200µL of ice-precooled mitochondrial separation solution (MSS) in a homogenizer with 20 times grinding. The homogenates were centrifuged at 800×g for 5min at 4°C to collect the nucleus at the bottom of the tube, and centrifuged at 8000×g for 10min at 4°C to collect the mitochondria. The supernatants were transferred to a new tube as the cytoplasmic component. The nucleus and the mitochondria pellets were then washed 3 times, and dissolved in 60µL of the NLB buffer [10mM Tris-HCl (pH7.5), 10mM NaCl, 3mM MgCl_2_, 0.3% (vol/vol) NP-40 and 10% (vol/vol) glycerol] or 100µL of the MSS buffer [10mM Tris-HCl (pH7.5), 250mM sucrose and 0.5mM EDTA], respectively. Finally, the nucleus and mitochondria were sonicated 3 times at 20% power for 15s in an ice bath with 2min of cooling between each sonication, followed by centrifuge at 12,000 rpm for 10min at 4°C. The protein concentration of each component was measured by the BCA kit. SAM contents in each subcellular component were measured using the ELISA kit (mlbio, China).

#### DNA Methylation

Mitochondrial and nuclear DNAs were extracted using a DNA extraction kit (ThermoFisher, #KIT0103) and quantified using NanoDrop (ThermoFisher). DNA methylation levels for each subcellular component were measured using the Global DNA methylation Detection Kit (Abcam, #ab233486) according to the manufacturer's instructions. In addition, the methylation of mtDNA in mice with *Slc25a26* overexpression measured by liquid chromatography-mass spectrometry (LC-MS). Briefly, 500ng mtRNAs were subjected to restriction enzyme digestion, then remove the potential bacterial contamination through ultrafiltration treatment. Afterwards, the mtDNA is dissolved in sterile water, heated at 95°C for 5min to denature, and then quickly transferred to ice for 2min. Following the addition of buffer solution, S1 nuclease, alkaline phosphatase, and DNase, the mtDNA was thoroughly hydrolyzed into nucleotides at 37°C. The sample after enzymatic digestion was extracted with chloroform and the upper aqueous solution was carefully collected. This collected solution, once concentrated, was reconstituted in ultrapure water, preparing it for subsequent LC-ESI-MS/MS analysis.

#### RNA Methylation

Cytoplasmic RNAs of the *Slc25a26*-flox and *Slc25a26*-cKO hearts were extracted as described above. RNA methylation levels were measured using the m^6^A RNA Methylation Analysis Kit (Abcam, #ab185912) and m^1^A Elisa Kit (Cell Biolabs, #MET-5099) according to the manufacturer’s instructions.

### SUPPLEMNETAL TABLES AND FIGURES

#### Supplemental Table S1. Slc25a26 deficiency modulates tRNA m1A modifications.

| **Fold enrichment** | | | | **Annotation** | | | | | | | |
| --- | --- | --- | --- | --- | --- | --- | --- | --- | --- | --- | --- |
| **Name** | **aver_flox** | **aver_cKO** | **Fold_change** | **name** | **chrom** | **txStart** | **txEnd** | **strand** | **isotype** | **anticodon** | **length** |
| tRNA-Ala-AGC-10-1 | 0.199697239 | 0.152726679 | 0.764791141 | chr13.trna1496 | chr13 | 23412940 | 23413013 | - | Ala | (AGC) | 73 |
| tRNA-Ala-AGC-1-1 | 0.187686773 | 0.085824848 | 0.45727702 | chr13.trna91 | chr13 | 21242569 | 21242641 | + | Ala | (AGC) | 72 |
| tRNA-Ala-AGC-2-1 | 0.068639451 | 0.03514811 | 0.512068636 | chr13.trna88 | chr13 | 21222585 | 21222657 | + | Ala | (AGC) | 72 |
| tRNA-Ala-AGC-3-1 | 0.818015066 | 0.417779569 | 0.510723563 | chr13.trna93 | chr13 | 21259767 | 21259839 | + | Ala | (AGC) | 72 |
| tRNA-Ala-AGC-4-1 | 0.05548909 | 0.009320059 | 0.167962014 | chr3.trna38 | chr3 | 19628994 | 19629067 | + | Ala | (AGC) | 73 |
| tRNA-Ala-AGC-5-1 | 0.106009021 | 0.182003947 | 1.716872259 | chr13.trna117 | chr13 | 23285648 | 23285721 | + | Ala | (AGC) | 73 |
| tRNA-Ala-AGC-6-1 | 0.016190088 | 0.035107959 | 2.168484693 | chr13.trna126 | chr13 | 23431411 | 23431484 | + | Ala | (AGC) | 73 |
| tRNA-Ala-AGC-7-1 | 0.021189726 | 0.026278056 | 1.240131937 | chr19.trna10 | chr19 | 3576262 | 3576335 | + | Ala | (AGC) | 73 |
| tRNA-Ala-AGC-8-1 | 2.799417576 | 4.069562932 | 1.453717719 | chr4.trna979 | chr4 | 132516007 | 132516079 | + | Ala | (AGC) | 72 |
| tRNA-Ala-CGC-1-2 | 3.797846098 | 2.570927928 | 0.676943684 | chr13.trna1488 | chr13 | 23438696 | 23438768 | - | Ala | (CGC) | 72 |
| tRNA-Ala-CGC-2-1 | 0.923967689 | 0.612685172 | 0.663102378 | chr2.trna412 | chr2 | 57182077 | 57182149 | + | Ala | (CGC) | 72 |
| tRNA-Ala-CGC-3-2 | 0.403796395 | 0.227717978 | 0.563942573 | chrX.trna1038 | chrX | 136395521 | 136395593 | - | Ala | (CGC) | 72 |
| tRNA-Ala-CGC-4-1 | 0.399296106 | 0.211801309 | 0.530436699 | chrX.trna602 | chrX | 136367720 | 136367792 | + | Ala | (CGC) | 72 |
| tRNA-Ala-CGC-5-1 | 1.74894207 | 4.476368475 | 2.559472125 | chr4.trna948 | chr4 | 132125258 | 132125330 | + | Ala | (CGC) | 72 |
| tRNA-Ala-CGC-6-1 | 0.717050067 | 0.413037897 | 0.57602379 | chrX.trna1035 | chrX | 136567864 | 136567936 | - | Ala | (CGC) | 72 |
| tRNA-Ala-CGC-7-1 | 0.423954789 | 0.179046273 | 0.422323977 | chrX.trna611 | chrX | 136569762 | 136569835 | + | Ala | (CGC) | 73 |
| tRNA-Ala-TGC-1-1 | 0.40115032 | 0.226710079 | 0.56514994 | chr13.trna90 | chr13 | 21240944 | 21241016 | + | Ala | (TGC) | 72 |
| tRNA-Ala-TGC-3-1 | 4.022062304 | 3.084150587 | 0.766808258 | chr11.trna3142 | chr11 | 48834428 | 48834500 | - | Ala | (TGC) | 72 |
| tRNA-Ala-TGC-4-1 | 1.713228847 | 1.226349041 | 0.715811576 | chr5.trna1075 | chr5 | 125409471 | 125409543 | + | Ala | (TGC) | 72 |
| tRNA-Ala-TGC-5-1 | 0.389664261 | 0.227593644 | 0.584076259 | chrX.trna604 | chrX | 136380746 | 136380818 | + | Ala | (TGC) | 72 |
| tRNA-Ala-TGC-6-1 | 0.413646884 | 0.236499833 | 0.571743295 | chrX.trna613 | chrX | 136590364 | 136590436 | + | Ala | (TGC) | 72 |
| tRNA-Ala-TGC-7-1 | 0.69858999 | 0.321381391 | 0.460042936 | chrX.trna1036 | chrX | 136430364 | 136430436 | - | Ala | (TGC) | 72 |
| tRNA-Ala-TGC-8-1 | 0.728977762 | 0.353636934 | 0.485113473 | chrX.trna601 | chrX | 136357601 | 136357673 | + | Ala | (TGC) | 72 |
| tRNA-Arg-ACG-1-2 | 0.344272945 | 0.533997469 | 1.551087521 | chr13.trna1484 | chr13 | 23500521 | 23500594 | - | Arg | (ACG) | 73 |
| tRNA-Arg-ACG-2-1 | 0.668384726 | 0.845714806 | 1.265311389 | chr3.trna1648 | chr3 | 19627958 | 19628031 | - | Arg | (ACG) | 73 |
| tRNA-Arg-ACG-3-2 | 0.427129075 | 0.679300652 | 1.590387289 | chr13.trna1507 | chr13 | 22021143 | 22021216 | - | Arg | (ACG) | 73 |
| tRNA-Arg-CCG-1-1 | 4.537594922 | 5.461844273 | 1.203687056 | chr13.trna1534 | chr13 | 21215457 | 21215530 | - | Arg | (CCG) | 73 |
| tRNA-Arg-CCG-2-1 | 29.85932242 | 19.82230769 | 0.663856581 | chr17.trna1621 | chr17 | 23550262 | 23550335 | - | Arg | (CCG) | 73 |
| tRNA-Arg-CCG-3-1 | 66.97375639 | 67.92300547 | 1.014173448 | chr11.trna1496 | chr11 | 107012865 | 107012938 | + | Arg | (CCG) | 73 |
| tRNA-Arg-CCT-1-1 | 1.388774237 | 1.540874917 | 1.10952153 | chr11.trna1594 | chr11 | 115412980 | 115413053 | + | Arg | (CCT) | 73 |
| tRNA-Arg-CCT-2-1 | 12.66865369 | 6.900155901 | 0.544663709 | chr11.trna1917 | chr11 | 115413303 | 115413376 | - | Arg | (CCT) | 73 |
| tRNA-Arg-CCT-3-1 | 3.619529711 | 2.954527562 | 0.81627388 | chr17.trna1622 | chr17 | 23548767 | 23548840 | - | Arg | (CCT) | 73 |
| tRNA-Arg-CCT-4-1 | 9.185238452 | 9.419270015 | 1.025479095 | chr6.trna174 | chr6 | 38533930 | 38534003 | + | Arg | (CCT) | 73 |
| tRNA-Arg-TCG-1-1 | 1.215258208 | 1.186247221 | 0.976127718 | chr7.trna629 | chr7 | 79466418 | 79466491 | + | Arg | (TCG) | 73 |
| tRNA-Arg-TCG-2-1 | 2.728072986 | 4.373488001 | 1.60314186 | chr11.trna1595 | chr11 | 115413778 | 115413851 | + | Arg | (TCG) | 73 |
| tRNA-Arg-TCG-3-2 | 4.227159986 | 4.735614727 | 1.120282824 | chr13.trna1482 | chr13 | 23503979 | 23504052 | - | Arg | (TCG) | 73 |
| tRNA-Arg-TCG-4-1 | 4.235815991 | 4.414501706 | 1.042184485 | chr13.trna1481 | chr13 | 23526455 | 23526528 | - | Arg | (TCG) | 73 |
| tRNA-Arg-TCT-1-1 | 0.049435688 | 1.730250461 | 35.00002823 | chr3.trna1019 | chr3 | 122292947 | 122293032 | - | Arg | (TCT) | 85 |
| tRNA-Arg-TCT-2-1 | 0.589786502 | 0.598061336 | 1.014030219 | chr11.trna2830 | chr11 | 69123481 | 69123568 | - | Arg | (TCT) | 87 |
| tRNA-Arg-TCT-3-1 | 0.03730297 | 0.121277184 | 3.25114017 | chr19.trna985 | chr19 | 12011587 | 12011673 | - | Arg | (TCT) | 86 |
| tRNA-Arg-TCT-5-1 | 0.178695864 | 0.047912424 | 0.268122736 | chr13.trna1519 | chr13 | 21917161 | 21917247 | - | Arg | (TCT) | 86 |
| tRNA-Asn-GTT-1-1 | 0.750361553 | 3.243570724 | 4.322677126 | chr3.trna1206 | chr3 | 96334614 | 96334688 | - | Asn | (GTT) | 74 |
| tRNA-Asn-GTT-2-1 | 0.079463395 | 0.093070498 | 1.171237365 | chr3.trna460 | chr3 | 96322073 | 96322147 | + | Asn | (GTT) | 74 |
| tRNA-Asn-GTT-3-1 | 0.70680793 | 1.264947178 | 1.789661835 | chr1.trna1458 | chr1 | 171033941 | 171034015 | - | Asn | (GTT) | 74 |
| tRNA-Asn-GTT-5-1 | 0.02 | 0.02 | 1 | chr3.trna462 | chr3 | 96334970 | 96335044 | + | Asn | (GTT) | 74 |
| tRNA-Asp-GTC-1-5 | 0.471601063 | 0.514707713 | 1.091404903 | chr1.trna1004 | chr1 | 171102370 | 171102442 | + | Asp | (GTC) | 72 |
| tRNA-Asp-GTC-2-1 | 0.661783683 | 1.648178275 | 2.490509086 | chr10.trna1337 | chr10 | 91182096 | 91182168 | - | Asp | (GTC) | 72 |
| tRNA-Asp-GTC-3-1 | 0.049794922 | 0.393208859 | 7.896565452 | chr2.trna794 | chr2 | 114103366 | 114103438 | + | Asp | (GTC) | 72 |
| tRNA-Asp-GTC-4-1 | 0.15780971 | 2.604783888 | 16.50585311 | chr5.trna2466 | chr5 | 100992873 | 100992945 | - | Asp | (GTC) | 72 |
| tRNA-Cys-GCA-10-1 | 2.339519162 | 11.19646234 | 4.785796381 | chr6.trna267 | chr6 | 48340612 | 48340684 | + | Cys | (GCA) | 72 |
| tRNA-Cys-GCA-11-1 | 5.181217869 | 7.697938988 | 1.485739296 | chr6.trna1587 | chr6 | 48288875 | 48288947 | - | Cys | (GCA) | 72 |
| tRNA-Cys-GCA-12-1 | 1.203480666 | 2.85174687 | 2.36958262 | chr6.trna1592 | chr6 | 48253079 | 48253151 | - | Cys | (GCA) | 72 |
| tRNA-Cys-GCA-13-1 | 1.152659128 | 1.845985404 | 1.601501571 | chr6.trna270 | chr6 | 48356850 | 48356922 | + | Cys | (GCA) | 72 |
| tRNA-Cys-GCA-14-1 | 2.011138763 | 2.379773893 | 1.183296716 | chr6.trna250 | chr6 | 48143355 | 48143427 | + | Cys | (GCA) | 72 |
| tRNA-Cys-GCA-15-1 | 1.163816835 | 1.658213371 | 1.424806139 | chr6.trna1583 | chr6 | 48308728 | 48308800 | - | Cys | (GCA) | 72 |
| tRNA-Cys-GCA-16-1 | 2.251832106 | 2.221482988 | 0.986522477 | chr6.trna1603 | chr6 | 48139509 | 48139581 | - | Cys | (GCA) | 72 |
| tRNA-Cys-GCA-17-1 | 2.025119728 | 2.36863934 | 1.169629285 | chr6.trna264 | chr6 | 48311498 | 48311570 | + | Cys | (GCA) | 72 |
| tRNA-Cys-GCA-18-1 | 1.154868686 | 1.733057003 | 1.500652865 | chr6.trna1584 | chr6 | 48305463 | 48305535 | - | Cys | (GCA) | 72 |
| tRNA-Cys-GCA-19-1 | 0.65666748 | 0.59935363 | 0.912720134 | chr6.trna255 | chr6 | 48186317 | 48186389 | + | Cys | (GCA) | 72 |
| tRNA-Cys-GCA-2-1 | 3.049910092 | 2.068692224 | 0.67827974 | chr11.trna2234 | chr11 | 97798905 | 97798977 | - | Cys | (GCA) | 72 |
| tRNA-Cys-GCA-21-1 | 2.09191807 | 2.533352556 | 1.211019013 | chr6.trna262 | chr6 | 48302458 | 48302530 | + | Cys | (GCA) | 72 |
| tRNA-Cys-GCA-24-1 | 0.21880732 | 2.585820701 | 11.81779794 | chr6.trna268 | chr6 | 48341587 | 48341659 | + | Cys | (GCA) | 72 |
| tRNA-Cys-GCA-25-1 | 0.207306753 | 0.7425449 | 3.58186547 | chr6.trna258 | chr6 | 48205067 | 48205139 | + | Cys | (GCA) | 72 |
| tRNA-Cys-GCA-3-2 | 1.990963325 | 2.520139119 | 1.26578882 | chr11.trna1235 | chr11 | 97797071 | 97797143 | + | Cys | (GCA) | 72 |
| tRNA-Cys-GCA-4-28 | 1.824365033 | 3.237661015 | 1.774678289 | chr6.trna1574 | chr6 | 48363531 | 48363603 | - | Cys | (GCA) | 72 |
| tRNA-Cys-GCA-5-1 | 2.074031428 | 0.919841716 | 0.443504232 | chr11.trna2236 | chr11 | 97769828 | 97769900 | - | Cys | (GCA) | 72 |
| tRNA-Cys-GCA-6-1 | 2.131890601 | 4.326816247 | 2.029567674 | chr6.trna1586 | chr6 | 48293006 | 48293078 | - | Cys | (GCA) | 72 |
| tRNA-Cys-GCA-7-1 | 0.819623029 | 1.053717439 | 1.285612289 | chr6.trna1598 | chr6 | 48209837 | 48209909 | - | Cys | (GCA) | 72 |
| tRNA-Cys-GCA-9-1 | 0.379524274 | 3.444975957 | 9.0770899 | chr6.trna263 | chr6 | 48303464 | 48303536 | + | Cys | (GCA) | 72 |
| tRNA-Gln-CTG-1-1 | 0.052695352 | 0.077220477 | 1.46541343 | chr13.trna110 | chr13 | 21920199 | 21920271 | + | Gln | (CTG) | 72 |
| tRNA-Gln-CTG-2-2 | 0.03254161 | 0.041464719 | 1.274206109 | chr11.trna2829 | chr11 | 69124606 | 69124678 | - | Gln | (CTG) | 72 |
| tRNA-Gln-CTG-3-2 | 0.054710166 | 0.065830099 | 1.203251684 | chr3.trna1201 | chr3 | 96399058 | 96399130 | - | Gln | (CTG) | 72 |
| tRNA-Gln-CTG-5-1 | 0.39734858 | 1.708882856 | 4.300714645 | chr13.trna1509 | chr13 | 21996580 | 21996652 | - | Gln | (CTG) | 72 |
| tRNA-Gln-TTG-1-1 | 0.051018024 | 0.060534301 | 1.186527743 | chr11.trna2317 | chr11 | 95859171 | 95859243 | - | Gln | (TTG) | 72 |
| tRNA-Gln-TTG-2-1 | 0.058852427 | 0.064215527 | 1.091127938 | chr13.trna1531 | chr13 | 21268468 | 21268540 | - | Gln | (TTG) | 72 |
| tRNA-Gln-TTG-3-1 | 0.053788744 | 0.062353734 | 1.159233863 | chr13.trna137 | chr13 | 23519049 | 23519121 | + | Gln | (TTG) | 72 |
| tRNA-Gln-TTG-5-1 | 0.053947742 | 0.060827869 | 1.127533188 | chr11.trna983 | chr11 | 86262167 | 86262239 | + | Gln | (TTG) | 72 |
| tRNA-Glu-CTC-1-4 | 8.398806363 | 9.792879892 | 1.165984721 | chr1.trna1006 | chr1 | 171103042 | 171103114 | + | Glu | (CTC) | 72 |
| tRNA-Glu-CTC-2-1 | 0.445188958 | 1.040695325 | 2.33764856 | chr17.trna1162 | chr17 | 56092000 | 56092072 | - | Glu | (CTC) | 72 |
| tRNA-Glu-CTC-5-1 | 3.82366952 | 4.837499662 | 1.26514586 | chr7.trna1598 | chr7 | 99543899 | 99543971 | - | Glu | (CTC) | 72 |
| tRNA-Glu-TTC-1-3 | 0.381701331 | 0.177460069 | 0.464918654 | chr13.trna129 | chr13 | 23463534 | 23463606 | + | Glu | (TTC) | 72 |
| tRNA-Glu-TTC-2-1 | 0.132513552 | 0.510935444 | 3.855722194 | chr1.trna2318 | chr1 | 34434811 | 34434883 | - | Glu | (TTC) | 72 |
| tRNA-Glu-TTC-3-2 | 1.900578389 | 1.258581203 | 0.662209573 | chr3.trna1202 | chr3 | 96387404 | 96387476 | - | Glu | (TTC) | 72 |
| tRNA-Gly-ACC-1-1 | 0.12889019 | 0.20005108 | 1.552104778 | chr11.trna324 | chr11 | 48823727 | 48823801 | + | Gly | (ACC) | 74 |
| tRNA-Gly-CCC-1-2 | 0.512549443 | 0.702741368 | 1.371070398 | chr17.trna174 | chr17 | 25875087 | 25875158 | + | Gly | (CCC) | 71 |
| tRNA-Gly-CCC-2-1 | 3.231056469 | 5.21649871 | 1.614487014 | chr3.trna1203 | chr3 | 96351166 | 96351237 | - | Gly | (CCC) | 71 |
| tRNA-Gly-CCC-3-1 | 3.940694389 | 4.788698498 | 1.215191544 | chr3.trna1200 | chr3 | 96424032 | 96424103 | - | Gly | (CCC) | 71 |
| tRNA-Gly-CCC-4-1 | 3.624545071 | 4.601579084 | 1.269560454 | chr4.trna96 | chr4 | 32230249 | 32230320 | + | Gly | (CCC) | 71 |
| tRNA-Gly-GCC-1-2 | 3.565280815 | 4.71276095 | 1.321848459 | chr1.trna1001 | chr1 | 171074301 | 171074372 | + | Gly | (GCC) | 71 |
| tRNA-Gly-GCC-2-1 | 5.265784928 | 5.593299009 | 1.062196631 | chr1.trna996 | chr1 | 171044984 | 171045055 | + | Gly | (GCC) | 71 |
| tRNA-Gly-GCC-3-1 | 6.186446508 | 7.448576704 | 1.204015374 | chr1.trna2073 | chr1 | 74816746 | 74816817 | - | Gly | (GCC) | 71 |
| tRNA-Gly-GCC-4-1 | 6.405598968 | 7.519963159 | 1.173967212 | chr8.trna854 | chr8 | 111062560 | 111062631 | + | Gly | (GCC) | 71 |
| tRNA-Gly-TCC-1-5 | 1.592633858 | 2.645674102 | 1.661194184 | chr1.trna1005 | chr1 | 171102755 | 171102827 | + | Gly | (TCC) | 72 |
| tRNA-His-GTG-1-1 | 0.04161663 | 0.020904577 | 0.502313061 | chr3.trna1197 | chr3 | 96463971 | 96464043 | - | His | (GTG) | 72 |
| tRNA-His-GTG-2-2 | 0.271763111 | 0.221911282 | 0.816561459 | chr2.trna2205 | chr2 | 122377362 | 122377434 | - | His | (GTG) | 72 |
| tRNA-His-GTG-3-1 | 0.339614818 | 0.265891046 | 0.782919451 | chr3.trna471 | chr3 | 96435971 | 96436043 | + | His | (GTG) | 72 |
| tRNA-Ile-AAT-1-2 | 0.042046951 | 0.122942618 | 2.92393656 | chr11.trna2835 | chr11 | 69062319 | 69062393 | - | Ile | (AAT) | 74 |
| tRNA-Ile-AAT-2-1 | 0.194762137 | 0.091511712 | 0.469863974 | chr4.trna87 | chr4 | 27904778 | 27904852 | + | Ile | (AAT) | 74 |
| tRNA-Ile-AAT-3-1 | 0.055288759 | 0.122768742 | 2.220500946 | chr13.trna123 | chr13 | 23397059 | 23397133 | + | Ile | (AAT) | 74 |
| tRNA-Ile-TAT-1-1 | 0.222649987 | 0.255461083 | 1.147366262 | chr7.trna259 | chr7 | 28372998 | 28373093 | + | Ile | (TAT) | 95 |
| tRNA-Ile-TAT-2-1 | 0.217895184 | 0.324558187 | 1.489515193 | chr13.trna1521 | chr13 | 21896937 | 21897031 | - | Ile | (TAT) | 94 |
| tRNA-Ile-TAT-2-2 | 0.228474659 | 0.205757932 | 0.900572223 | chr13.trna1502 | chr13 | 23270420 | 23270516 | - | Ile | (TAT) | 96 |
| tRNA-Ile-TAT-2-3 | 0.355078772 | 0.375219276 | 1.056721229 | chr17.trna803 | chr17 | 83861943 | 83862036 | + | Ile | (TAT) | 93 |
| tRNA-Ile-TAT-3-1 | 0.02 | 0.113202368 | 5.660118398 | chr1.trna838 | chr1 | 153458663 | 153458736 | + | Ile | (TAT) | 73 |
| tRNA-iMet-CAT-1-2 | 0.369981477 | 0.919260691 | 2.484612735 | chr13.trna101 | chr13 | 21710993 | 21711065 | + | iMet | (CAT) | 72 |
| tRNA-iMet-CAT-2-1 | 0.285324914 | 0.685804713 | 2.403592111 | chr15.trna1319 | chr15 | 69396622 | 69396694 | - | iMet | (CAT) | 72 |
| tRNA-Leu-AAG-1-1 | 4.268961648 | 3.041249379 | 0.712409628 | chr11.trna3141 | chr11 | 48856973 | 48857055 | - | Leu | (AAG) | 82 |
| tRNA-Leu-AAG-2-1 | 4.197967766 | 2.855582091 | 0.680229637 | chr7.trna917 | chr7 | 120917516 | 120917598 | + | Leu | (AAG) | 82 |
| tRNA-Leu-AAG-3-1 | 2.555301512 | 3.663889305 | 1.43383835 | chr13.trna85 | chr13 | 21170205 | 21170287 | + | Leu | (AAG) | 82 |
| tRNA-Leu-CAA-1-1 | 8.958527172 | 7.825672543 | 0.873544545 | chr13.trna1536 | chr13 | 21172836 | 21172942 | - | Leu | (CAA) | 106 |
| tRNA-Leu-CAA-2-1 | 1.818325429 | 1.784116484 | 0.981186567 | chr13.trna87 | chr13 | 21200203 | 21200308 | + | Leu | (CAA) | 105 |
| tRNA-Leu-CAA-3-1 | 1.916955115 | 1.907877688 | 0.995264664 | chr11.trna2968 | chr11 | 58302473 | 58302580 | - | Leu | (CAA) | 107 |
| tRNA-Leu-CAA-4-1 | 1.889466321 | 2.319156729 | 1.227413637 | chr13.trna108 | chr13 | 21914837 | 21914943 | + | Leu | (CAA) | 106 |
| tRNA-Leu-CAG-1-2 | 2.725922026 | 2.327858681 | 0.853971118 | chr1.trna1000 | chr1 | 171073618 | 171073701 | + | Leu | (CAG) | 83 |
| tRNA-Leu-CAG-2-1 | 11.52866816 | 8.529427852 | 0.739845031 | chr1.trna995 | chr1 | 171038613 | 171038696 | + | Leu | (CAG) | 83 |
| tRNA-Leu-CAG-3-1 | 2.614010366 | 2.149830409 | 0.822426122 | chr8.trna261 | chr8 | 38213200 | 38213283 | + | Leu | (CAG) | 83 |
| tRNA-Leu-CAG-4-1 | 1.693097854 | 1.788773382 | 1.056509154 | chr3.trna1632 | chr3 | 24163024 | 24163107 | - | Leu | (CAG) | 83 |
| tRNA-Leu-TAA-1-1 | 0.22065788 | 0.970302866 | 4.397317983 | chr19.trna986 | chr19 | 12011085 | 12011168 | - | Leu | (TAA) | 83 |
| tRNA-Leu-TAA-2-1 | 0.460520153 | 0.342376604 | 0.743456288 | chr10.trna2054 | chr10 | 12922570 | 12922653 | - | Leu | (TAA) | 83 |
| tRNA-Leu-TAG-1-1 | 1.390672807 | 0.843557997 | 0.60658265 | chr11.trna623 | chr11 | 69124240 | 69124322 | + | Leu | (TAG) | 82 |
| tRNA-Leu-TAG-2-1 | 2.0604276 | 1.731665983 | 0.840440102 | chr14.trna1079 | chr14 | 51082829 | 51082911 | - | Leu | (TAG) | 82 |
| tRNA-Leu-TAG-3-1 | 1.153387117 | 0.7080151 | 0.613857298 | chr7.trna1422 | chr7 | 120835478 | 120835560 | - | Leu | (TAG) | 82 |
| tRNA-Lys-CTT-1-1 | 0.117688093 | 0.03741554 | 0.317921206 | chrX.trna1043 | chrX | 136195731 | 136195804 | - | Lys | (CTT) | 73 |
| tRNA-Lys-CTT-12-1 | 0.584815538 | 0.02 | 0.034198818 | chr3.trna1 | chr3 | 3123739 | 3123812 | + | Lys | (CTT) | 73 |
| tRNA-Lys-CTT-13-1 | 0.28109762 | 0.639528703 | 2.275112478 | chr2.trna2489 | chr2 | 86140983 | 86141060 | - | Lys | (CTT) | 77 |
| tRNA-Lys-CTT-14-1 | 0.123163434 | 0.042064307 | 0.341532434 | chr16.trna440 | chr16 | 47250545 | 47250618 | + | Lys | (CTT) | 73 |
| tRNA-Lys-CTT-2-2 | 0.511787258 | 0.78665149 | 1.53706736 | chr12.trna1324 | chr12 | 70969119 | 70969192 | - | Lys | (CTT) | 73 |
| tRNA-Lys-CTT-3-3 | 1.30911009 | 1.307651963 | 0.998886169 | chr11.trna3143 | chr11 | 48833882 | 48833955 | - | Lys | (CTT) | 73 |
| tRNA-Lys-CTT-4-1 | 0.182793939 | 0.200253655 | 1.095515837 | chr3.trna2 | chr3 | 3149492 | 3149565 | + | Lys | (CTT) | 73 |
| tRNA-Lys-CTT-5-1 | 0.250646457 | 4.24575601 | 16.93922214 | chr5.trna2493 | chr5 | 98041089 | 98041162 | - | Lys | (CTT) | 73 |
| tRNA-Lys-CTT-8-1 | 0.298161305 | 0.320364243 | 1.074466197 | chr16.trna122 | chr16 | 16854263 | 16854336 | + | Lys | (CTT) | 73 |
| tRNA-Lys-CTT-9-1 | 0.527333984 | 0.825117117 | 1.56469551 | chr11.trna2738 | chr11 | 74137314 | 74137388 | - | Lys | (CTT) | 74 |
| tRNA-Lys-TTT-1-2 | 0.079939798 | 0.048113289 | 0.601869037 | chr1.trna1742 | chr1 | 133034326 | 133034399 | - | Lys | (TTT) | 73 |
| tRNA-Lys-TTT-2-2 | 0.022339359 | 0.063233923 | 2.830605956 | chr13.trna106 | chr13 | 21907939 | 21908012 | + | Lys | (TTT) | 73 |
| tRNA-Lys-TTT-3-1 | 0.02 | 0.02 | 1 | chr13.trna112 | chr13 | 21971787 | 21971860 | + | Lys | (TTT) | 73 |
| tRNA-Lys-TTT-5-1 | 0.115765936 | 0.02 | 0.172762392 | chr10.trna1378 | chr10 | 87829885 | 87829958 | - | Lys | (TTT) | 73 |
| tRNA-Met-CAT-1-2 | 1.637107507 | 1.086819107 | 0.663865447 | chr15.trna1375 | chr15 | 57975982 | 57976055 | - | Met | (CAT) | 73 |
| tRNA-Met-CAT-2-2 | 5.659782349 | 5.274176354 | 0.931869112 | chr13.trna1537 | chr13 | 21169686 | 21169759 | - | Met | (CAT) | 73 |
| tRNA-Met-CAT-3-1 | 1.635856405 | 1.456095023 | 0.89011176 | chr13.trna1530 | chr13 | 21326716 | 21326789 | - | Met | (CAT) | 73 |
| tRNA-Met-CAT-4-1 | 0.950266826 | 0.608781007 | 0.640642176 | chr13.trna122 | chr13 | 23317465 | 23317538 | + | Met | (CAT) | 73 |
| tRNA-Met-CAT-5-1 | 0.226361209 | 0.098426009 | 0.434818354 | chr15.trna550 | chr15 | 88641531 | 88641604 | + | Met | (CAT) | 73 |
| tRNA-Phe-GAA-1-2 | 3.741358355 | 3.779552124 | 1.01020853 | chr10.trna1481 | chr10 | 80248963 | 80249036 | - | Phe | (GAA) | 73 |
| tRNA-Phe-GAA-2-1 | 4.119826589 | 5.282036492 | 1.282101656 | chr19.trna174 | chr19 | 12008544 | 12008617 | + | Phe | (GAA) | 73 |
| tRNA-Phe-GAA-3-1 | 0.574820051 | 0.433380782 | 0.753941657 | chr13.trna1522 | chr13 | 21882884 | 21882957 | - | Phe | (GAA) | 73 |
| tRNA-Pro-AGG-1-1 | 1.809962303 | 1.570415968 | 0.867651202 | chr1.trna958 | chr1 | 165641633 | 165641705 | + | Pro | (AGG) | 72 |
| tRNA-Pro-CGG-1-1 | 1.854746608 | 1.619916231 | 0.87338951 | chr1.trna1491 | chr1 | 165642270 | 165642342 | - | Pro | (CGG) | 72 |
| tRNA-Pro-TGG-1-1 | 1.706222546 | 1.49635542 | 0.876998972 | chr7.trna725 | chr7 | 98815414 | 98815486 | + | Pro | (TGG) | 72 |
| tRNA-Pro-TGG-2-2 | 1.911189784 | 1.669574072 | 0.873578378 | chr11.trna327 | chr11 | 48856279 | 48856351 | + | Pro | (TGG) | 72 |
| tRNA-Pro-TGG-3-1 | 1.934827755 | 1.816603249 | 0.938896625 | chr11.trna585 | chr11 | 67291138 | 67291210 | + | Pro | (TGG) | 72 |
| tRNA-SeC-TCA-1-1 | 1.960928826 | 4.569348204 | 2.330195845 | chr7.trna147 | chr7 | 19301243 | 19301330 | + | SeC | (TCA) | 87 |
| tRNA-Ser-AGA-1-1 | 0.910375091 | 1.889055492 | 2.075029855 | chr13.trna1514 | chr13 | 21937093 | 21937175 | - | Ser | (AGA) | 82 |
| tRNA-Ser-AGA-2-2 | 0.729430468 | 1.381533855 | 1.893989786 | chr11.trna616 | chr11 | 69037476 | 69037558 | + | Ser | (AGA) | 82 |
| tRNA-Ser-CGA-1-1 | 0.099814378 | 0.214248363 | 2.146467939 | chr11.trna622 | chr11 | 69110918 | 69111000 | + | Ser | (CGA) | 82 |
| tRNA-Ser-CGA-2-1 | 0.099695334 | 0.215115503 | 2.157728887 | chr10.trna1048 | chr10 | 128458463 | 128458545 | - | Ser | (CGA) | 82 |
| tRNA-Ser-CGA-3-1 | 0.02 | 0.024114582 | 1.205729106 | chr13.trna1506 | chr13 | 22022418 | 22022500 | - | Ser | (CGA) | 82 |
| tRNA-Ser-GCT-1-1 | 7.997356105 | 8.669685206 | 1.084068921 | chr13.trna1510 | chr13 | 21994189 | 21994271 | - | Ser | (GCT) | 82 |
| tRNA-Ser-GCT-2-1 | 7.376625543 | 8.984263761 | 1.217936807 | chr2.trna2328 | chr2 | 119046749 | 119046831 | - | Ser | (GCT) | 82 |
| tRNA-Ser-GCT-3-1 | 0.958111945 | 1.682612337 | 1.756175096 | chr19.trna1108 | chr19 | 5038303 | 5038385 | - | Ser | (GCT) | 82 |
| tRNA-Ser-GCT-4-1 | 16.88823933 | 19.72935496 | 1.168230422 | chr11.trna2833 | chr11 | 69063041 | 69063123 | - | Ser | (GCT) | 82 |
| tRNA-Ser-GCT-5-1 | 10.30543777 | 19.29271072 | 1.872090361 | chr13.trna1527 | chr13 | 21492844 | 21492926 | - | Ser | (GCT) | 82 |
| tRNA-Ser-GGA-1-1 | 0.327382906 | 0.26622271 | 0.813184515 | chr11.trna1301 | chr11 | 100538157 | 100538230 | + | Ser | (GGA) | 73 |
| tRNA-Ser-TGA-1-1 | 0.375378358 | 0.686315067 | 1.828328811 | chr10.trna1702 | chr10 | 63429475 | 63429557 | - | Ser | (TGA) | 82 |
| tRNA-Ser-TGA-2-2 | 0.945815098 | 1.836151896 | 1.941343397 | chr13.trna136 | chr13 | 23518145 | 23518227 | + | Ser | (TGA) | 82 |
| tRNA-Sup-TCA-1-1 | 0.268666992 | 0.02 | 0.074441597 | chr5.trna18 | chr5 | 5627680 | 5627751 | + | Sup | (TCA) | 71 |
| tRNA-Thr-AGT-1-3 | 0.242946152 | 0.33120084 | 1.363268519 | chr11.trna2834 | chr11 | 69062743 | 69062817 | - | Thr | (AGT) | 74 |
| tRNA-Thr-AGT-2-1 | 0.100508086 | 0.33379028 | 3.321029117 | chr13.trna128 | chr13 | 23450621 | 23450695 | + | Thr | (AGT) | 74 |
| tRNA-Thr-AGT-3-1 | 0.23027777 | 0.330397638 | 1.434778695 | chr11.trna621 | chr11 | 69110464 | 69110538 | + | Thr | (AGT) | 74 |
| tRNA-Thr-AGT-4-1 | 0.042297674 | 0.171698417 | 4.059287458 | chr13.trna104 | chr13 | 21856020 | 21856094 | + | Thr | (AGT) | 74 |
| tRNA-Thr-AGT-5-1 | 0.059426814 | 0.035025249 | 0.589384609 | chr13.trna1505 | chr13 | 22030099 | 22030173 | - | Thr | (AGT) | 74 |
| tRNA-Thr-AGT-6-1 | 0.02 | 0.869869898 | 43.4934949 | chr13.trna1532 | chr13 | 21249193 | 21249267 | - | Thr | (AGT) | 74 |
| tRNA-Thr-AGT-7-1 | 0.788548889 | 1.13722276 | 1.442171532 | chr14.trna970 | chr14 | 58818152 | 58818226 | - | Thr | (AGT) | 74 |
| tRNA-Thr-CGT-1-1 | 1.56893498 | 1.715611084 | 1.093487688 | chr16.trna90 | chr16 | 13437322 | 13437394 | + | Thr | (CGT) | 72 |
| tRNA-Thr-CGT-2-1 | 2.194956265 | 1.967386539 | 0.896321522 | chr11.trna860 | chr11 | 79704649 | 79704721 | + | Thr | (CGT) | 72 |
| tRNA-Thr-CGT-3-1 | 0.317936895 | 0.072888943 | 0.229256007 | chr13.trna97 | chr13 | 21322812 | 21322886 | + | Thr | (CGT) | 74 |
| tRNA-Thr-CGT-4-1 | 0.169452255 | 0.088498631 | 0.522262929 | chr13.trna94 | chr13 | 21264579 | 21264653 | + | Thr | (CGT) | 74 |
| tRNA-Thr-TGT-1-1 | 0.143505544 | 1.066413045 | 7.431162682 | chr13.trna95 | chr13 | 21275955 | 21276029 | + | Thr | (TGT) | 74 |
| tRNA-Thr-TGT-2-1 | 0.70301432 | 1.003781818 | 1.427825564 | chr14.trna292 | chr14 | 51088649 | 51088722 | + | Thr | (TGT) | 73 |
| tRNA-Thr-TGT-3-1 | 0.740793 | 0.990613802 | 1.337234291 | chr11.trna326 | chr11 | 48853618 | 48853691 | + | Thr | (TGT) | 73 |
| tRNA-Trp-CCA-1-1 | 3.620521428 | 2.739955579 | 0.756784798 | chr11.trna2879 | chr11 | 62746014 | 62746086 | - | Trp | (CCA) | 72 |
| tRNA-Trp-CCA-2-1 | 56.0867827 | 35.11710954 | 0.626120948 | chr10.trna1338 | chr10 | 91181411 | 91181483 | - | Trp | (CCA) | 72 |
| tRNA-Trp-CCA-3-2 | 3.703867652 | 2.7674998 | 0.747191871 | chr11.trna2832 | chr11 | 69063494 | 69063566 | - | Trp | (CCA) | 72 |
| tRNA-Trp-CCA-4-1 | 3.939303657 | 2.917240027 | 0.740547132 | chr13.trna131 | chr13 | 23498105 | 23498177 | + | Trp | (CCA) | 72 |
| tRNA-Trp-CCA-5-1 | 2.45518419 | 2.094568571 | 0.853120748 | chr11.trna2922 | chr11 | 61407915 | 61407987 | - | Trp | (CCA) | 72 |
| tRNA-Trp-CCA-6-1 | 3.633232621 | 2.814255206 | 0.774587124 | chr10.trna2003 | chr10 | 23777082 | 23777154 | - | Trp | (CCA) | 72 |
| tRNA-Tyr-GTA-1-2 | 0.139795732 | 0.117996564 | 0.844064136 | chr13.trna125 | chr13 | 23426504 | 23426592 | + | Tyr | (GTA) | 88 |
| tRNA-Tyr-GTA-1-3 | 0.151076351 | 0.11510668 | 0.761910647 | chr13.trna1493 | chr13 | 23427094 | 23427180 | - | Tyr | (GTA) | 86 |
| tRNA-Tyr-GTA-1-5 | 0.149646496 | 0.132846149 | 0.887733107 | chr13.trna130 | chr13 | 23467075 | 23467165 | + | Tyr | (GTA) | 90 |
| tRNA-Tyr-GTA-3-1 | 0.112751364 | 0.115325408 | 1.022829385 | chr3.trna37 | chr3 | 19628781 | 19628870 | + | Tyr | (GTA) | 89 |
| tRNA-Tyr-GTA-3-2 | 0.115711329 | 0.109909591 | 0.949860241 | chr14.trna293 | chr14 | 51089488 | 51089578 | + | Tyr | (GTA) | 90 |
| tRNA-Tyr-GTA-4-1 | 0.106048202 | 0.086564857 | 0.816278401 | chr3.trna36 | chr3 | 19628354 | 19628447 | + | Tyr | (GTA) | 93 |
| tRNA-Tyr-GTA-5-1 | 0.1071283 | 0.102569439 | 0.957444845 | chr13.trna1492 | chr13 | 23428737 | 23428826 | - | Tyr | (GTA) | 89 |
| tRNA-Val-AAC-1-2 | 0.196317167 | 0.339494136 | 1.729314559 | chr11.trna328 | chr11 | 48856636 | 48856709 | + | Val | (AAC) | 73 |
| tRNA-Val-AAC-2-1 | 0.130927816 | 0.201995409 | 1.54279981 | chr13.trna116 | chr13 | 23285400 | 23285473 | + | Val | (AAC) | 73 |
| tRNA-Val-AAC-3-1 | 0.308657006 | 0.281241502 | 0.911178094 | chr13.trna1498 | chr13 | 23401072 | 23401145 | - | Val | (AAC) | 73 |
| tRNA-Val-AAC-4-1 | 0.277600561 | 0.366689888 | 1.320926321 | chr13.trna118 | chr13 | 23298780 | 23298853 | + | Val | (AAC) | 73 |
| tRNA-Val-AAC-5-1 | 0.189885538 | 0.324831994 | 1.710672641 | chr13.trna1508 | chr13 | 22015790 | 22015863 | - | Val | (AAC) | 73 |
| tRNA-Val-CAC-1-1 | 0.190812285 | 0.325506673 | 1.705899976 | chr13.trna121 | chr13 | 23307628 | 23307701 | + | Val | (CAC) | 73 |
| tRNA-Val-CAC-2-1 | 0.224351124 | 0.351556409 | 1.56699197 | chr1.trna1007 | chr1 | 171111442 | 171111515 | + | Val | (CAC) | 73 |
| tRNA-Val-CAC-3-1 | 0.411973229 | 0.287272445 | 0.697308526 | chr13.trna120 | chr13 | 23300121 | 23300194 | + | Val | (CAC) | 73 |
| tRNA-Val-CAC-5-1 | 0.02 | 0.354684964 | 17.7342482 | chr13.trna1497 | chr13 | 23412076 | 23412149 | - | Val | (CAC) | 73 |
| tRNA-Val-CAC-7-1 | 0.02 | 0.116759504 | 5.837975224 | chr3.trna1488 | chr3 | 59474925 | 59474997 | - | Val | (CAC) | 72 |
| tRNA-Val-TAC-1-1 | 0.070727737 | 0.232884808 | 3.292694207 | chr19.trna175 | chr19 | 12011930 | 12012003 | + | Val | (TAC) | 73 |

#### Supplemental Table S2.

|  | Gene | Species | Sequence (sense/forward) | Sequence (antisense/reverse) |
| --- | --- | --- | --- | --- |
| Primers |  |  |  |  |
| m_Slc25a26 | Slc25a26 | Mouse | GAAGAGGGCATTCAAGGACTGT | CAGAGGGCTTTCAAGGATTCCCA |
| m_Nppa | Nppa | Mouse | TTTCAAGAACCTGCTAGACCACC | GATCTATCGGAGGGGTCCCA |
| m_Nppb | Nppb | Mouse | CGCTGGGAGGTCACTCCTAT | CTTCAGTGCGTTACAGCCCAA |
| m_Myh6 | Myh6 | Mouse | AGCTCACCTACCAGACAGAGG | TTCCTCGTCGTGCATCTTCTT |
| m_Myh7 | Myh7 | Mouse | GGCCTGGGCTTACCTCTCTA | ACAGTCACCGTCTTGCCATT |
| m_Trmt61a | Trmt61a | Mouse | AGGAATTCCAGGAGCATCGG | AGAAACGCCCACCTTCAACC |
| m_Cytb | Cytb | Mouse | ACCTCCTATCAGCCATCCCA | GATTGCTAGGGCCGCGATAA |
| m_ND1 | ND1 | Mouse | CAACCATTTGCAGACGCCAT | GGGTGTGGTATTGGTAGGGG |
| m_H19 | H19 | Mouse | AGTGCCTCATGGGAATGGTG | GTGTCACCAGAAGGGGAGTG |
| m_GAPDH | GAPDH | Mouse | TCCTGCACCACCAACTGCTTAG | GATGACCTTGCCCACAGCCTTG |
| r_Slc25a26 | Slc25a26 | Rat | CTGGCGGCTTTCGTGGAATA | CTGGCGGCTTTCGTGGAATA |
| r_Nppa | Nppa | Rat | ATACAGTGCGGTGTCCAACA | AGCCCTCAGTTTGCTTTTCA |
| r_Nppb | Nppb | Rat | CAGCTCTCAAAGGACCAAGG | GCAGCTTGAACTATGTGCCA |
| r_Myh6 | Myh6 | Rat | ACTCATGGCCACACTCTTCT | AAGTGAGGATGGGTGGTCCT |
| r_Myh7 | Myh7 | Rat | GCTCCTAAGTAATCTGTTTG | AAGTGAGGGTGCGTGGAGCG |
| r_Trmt6 | Trmt6 | Rat | AGAGCCCACATCAGAGACCA | GCCCTTGTCTTTCAGAGCCT |
| r_Trmt61a | Trmt61a | Rat | GGTTTATGCTGCGTCCCTGA | CAGTGACAGGATGGCTGTGT |
| r_Alkbh1 | Alkbh1 | Rat | TTCCGAGCAGAAGCAGGTAT | GTGCATAAACATGGCTGTGG |
| r_Alkbh3 | Alkbh3 | Rat | CATGGTCTGAATGGGGAACGA | GCTTCTAGCAGGAGTAGCTGG |
| r_GAPDH | GAPDH | Rat | ACAGCAACAGGGTGGTGGAC | TTTGAGGGTGCAGCGAACTT |
| siRNAs/shRNAs |  |  |  |  |
| siSlc25a26-Mus | Slc25a26 | Mouse | CGUCUCUGUGGACUUGAUAUUAUTT | AAAUAAUAUCAAGUCCACAGAGACGCC |
| siSlc25a26-Rat | Slc25a26 | Rat | GGACUUGAUAUUAUUUCCUUUGGAT | AUCCAAAGGAAAUAAUAUCAAGUCCAC |
| siTrmt6-Rat | Trmt6 | Rat | CGAAUGGGAGGCUUUGGCUCCAUUA | UAAUGGAGCCAAAGCCUCCCAUUCG |
| siTrmt61a-Rat | Trmt61a | Rat | CACCGCACACAGAUCCUCUACUCUA | UAGAGUAGAGGAUCUGUGUGCGGUG |
| siAlkbh1-Rat | Alkbh1 | Rat | CAGUAAGAAAUACUCAGCAGAUCAU | AUGAUCUGCUGAGUAUUUCUUACUG |
| siAlkbh3-Rat | Alkbh3 | Rat | GAAUGAGAGCUACCAACACUUCATG | CAUGAAGUGUUGGUAGCUCUCAUUCAG |
| shTrmt61a-Mus | Trmt61a | Mouse | GGAGGTGATGATAATCAATAATCAT | ATGATTATTGATTATCATCACCTCC |

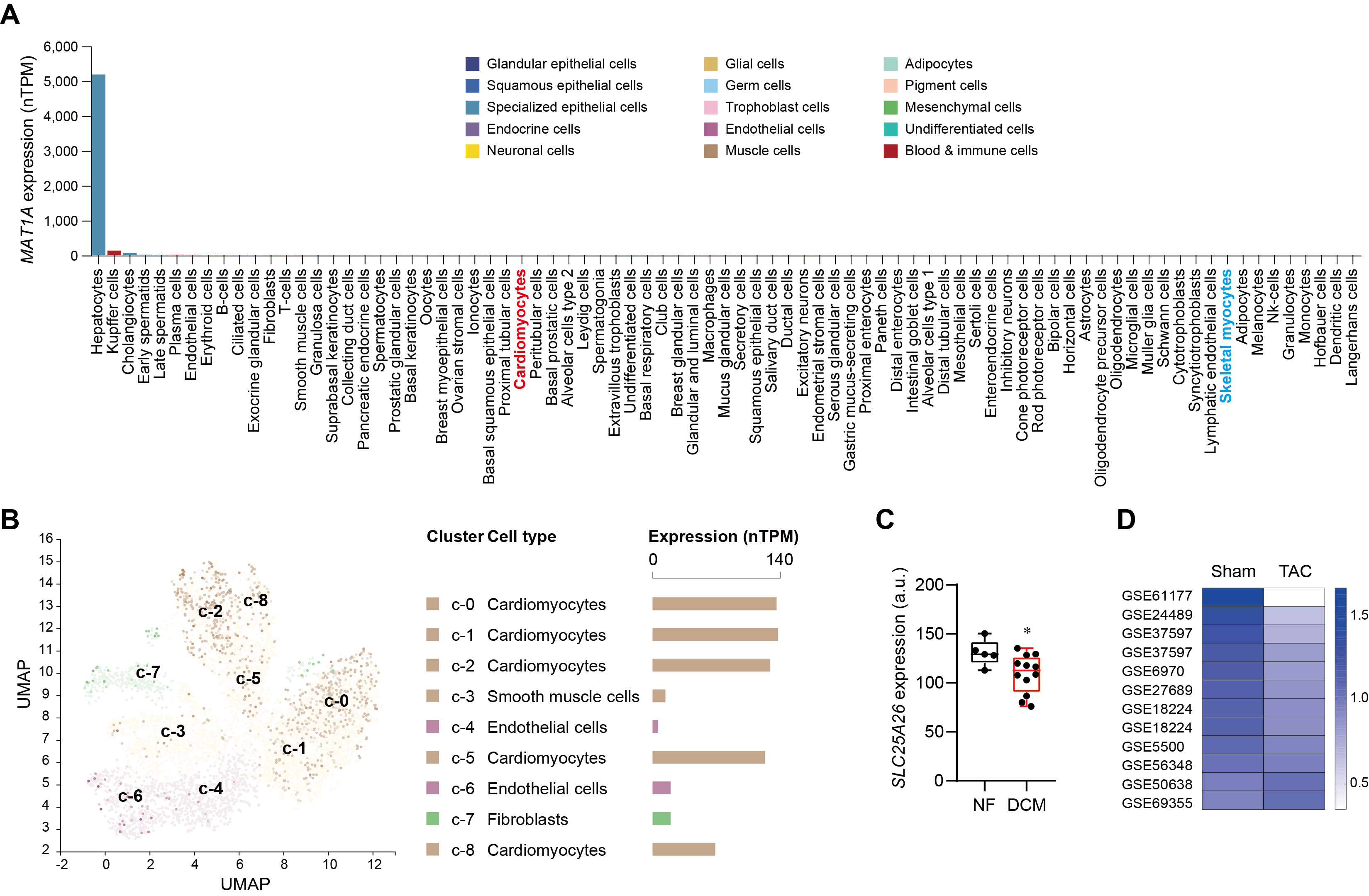

### Figure S1. Cell-type-specific expression of *MAT1A* and *SLC25a26*, and the alterations of SLC25A26 in human dilated cardiomyopathy and mouse cardiac hypertrophy induced by transaortic constriction (TAC).

**A** and **B**, Cell-type specific expression of *MAT1A* (**A**) and *SLC25A26* in the heart (**B**) from a single-cell sequencing database. Data are from the Human Protein Atlas (<https://www.proteinatlas.org/>).

**C**, *SLC25A26* expression in human hearts from patients with dilated cardiomyopathy (DCM) with non-failing (NF) hearts as the control. *N* = 5 (NF) and 12 (DCM). Data are from Gene Expression Omnibus (GEO) datasets (Accession: GSE42955).

**D**, Heatmap showing the alteration of *Slc25a26* expression in public available TAC datasets with GEO accession numbers labeled on the figure.

Data were presented as mean ± SEM of at least three independent experiments, unless denoted elsewhere. *p < 0.05, **p < 0.01, ***p < 0.001 vs. NF, (**C**, unpaired Student’s t test).

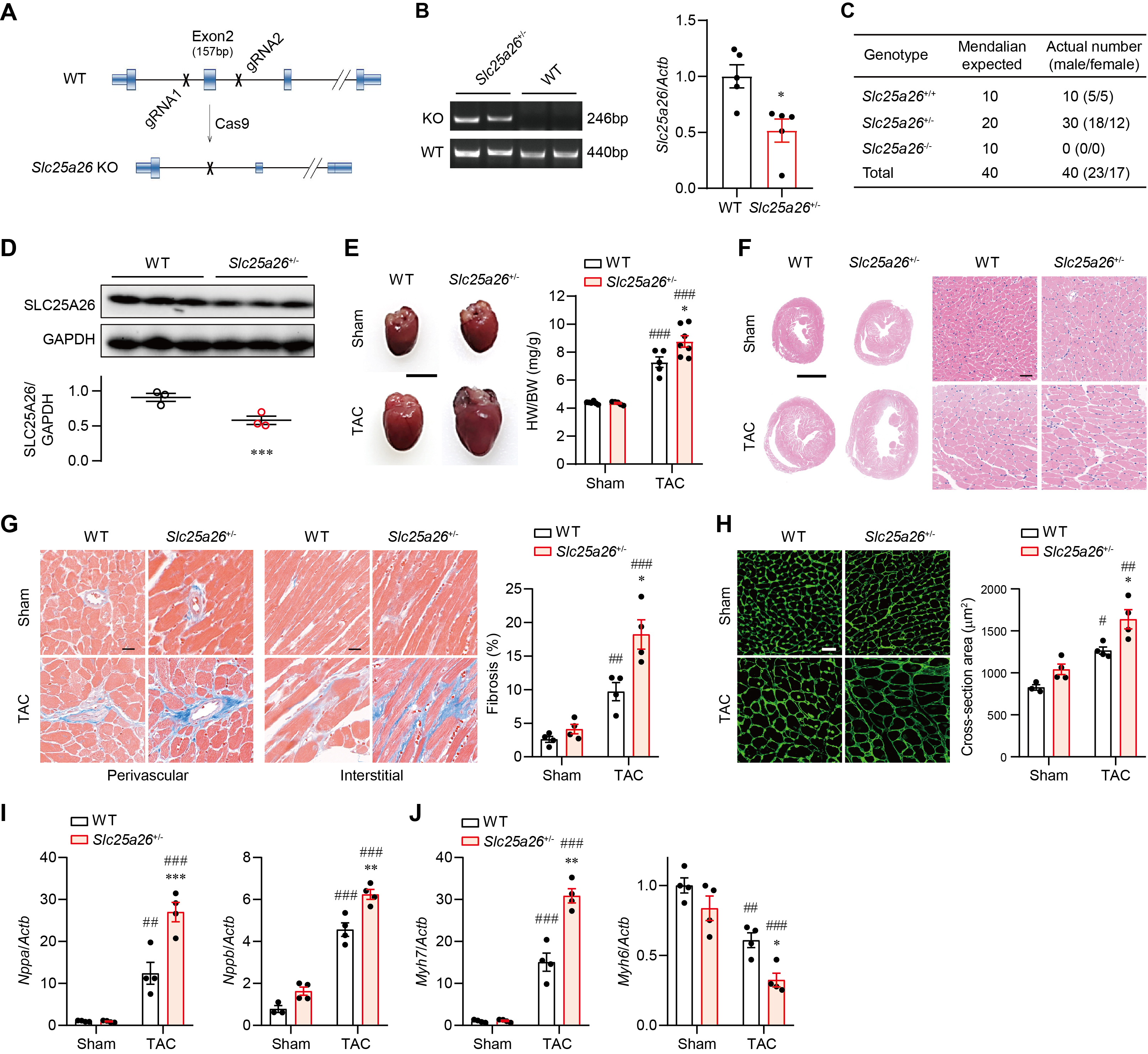

### Figure S2. Monoallelic loss of *Slc25a26* aggravates TAC-induced cardiac hypertrophy.

**A**, Schematic illustrating the *Slc25a26* knockout strategy.

**B**, Genotyping validation of *Slc25a26* knockout by RT-PCR (left) and quantification of *Slc25a26* expression by qRT-PCR (right) in wildtype (WT; *Slc25a26*^+/+^) and heterozygous Slc25a26 knockout (*Slc25a26*^+/-^) hearts. *N* = 5.

**C**, Genotyping quantification of the offspring from intercrosses of *Slc25a26*^+/-^ mice showing that biallelic *Slc25a26* knockout (*Slc25a26*^-/-^) was embryonic lethal.

**D**, *Slc25a26* expression at the protein levels in *Slc25a26*^+/+^ and *Slc25a26*^+/-^ hearts measured by Western blot. *N* = 3.

**E**, Representative morphology images (left) and weights (right) of WT and *Slc25a26*^+/-^ hearts with sham or TAC surgeries. HW, heart weight; BW, body weight. Scale bar, 5mm. *N* = 5~7 as indicated by dots on each bar.

**F**, Representative low- (left) and high-magnification (right) H&E-stained images of WT and *Slc25a26*^+/-^ hearts with sham or TAC surgeries. Scale bar, 2mm (left) and 20μm (right).

**G**, Representative Masson trichrome staining images (left and middle) and quantification (right) showing the impact of monoallelic *Slc25a26* deletion on myocardial fibrosis. Scale bar, 20μm. *N* = 4.

**H**, Representative wheat germ agglutinin (WGA) staining images (left) and quantification (right) showing the impact of monoallelic *Slc25a26* deletion on cross-sectional area of cardiomyocytes. Scale bar, 20μm. *N* = 4.

**I** and **J**, Impact of monoallelic *Slc25a26* deletion on TAC-induced expressions of hypertrophic biomarkers natriuretic peptide A (Nppa) and natriuretic peptide B (Nppb) (**I**) and the fetal gene reprogramming from myosin heavy chain (Myh) 6 to Myh7 (**J**). *N* = 4.

Data were presented as mean ± SEM of at least three independent experiments, unless denoted elsewhere. *p < 0.05, **p < 0.01, ***p < 0.001 vs. WT; ^#^*P* < 0.05, ^##^*P* < 0.01, ^###^*P* < 0.001 vs. sham, (**B** and **D**, unpaired Student’s t test; **E**, **G**, **H**, **I** and **J**, two-way ANOVA with Tukey’s multiple comparison test).

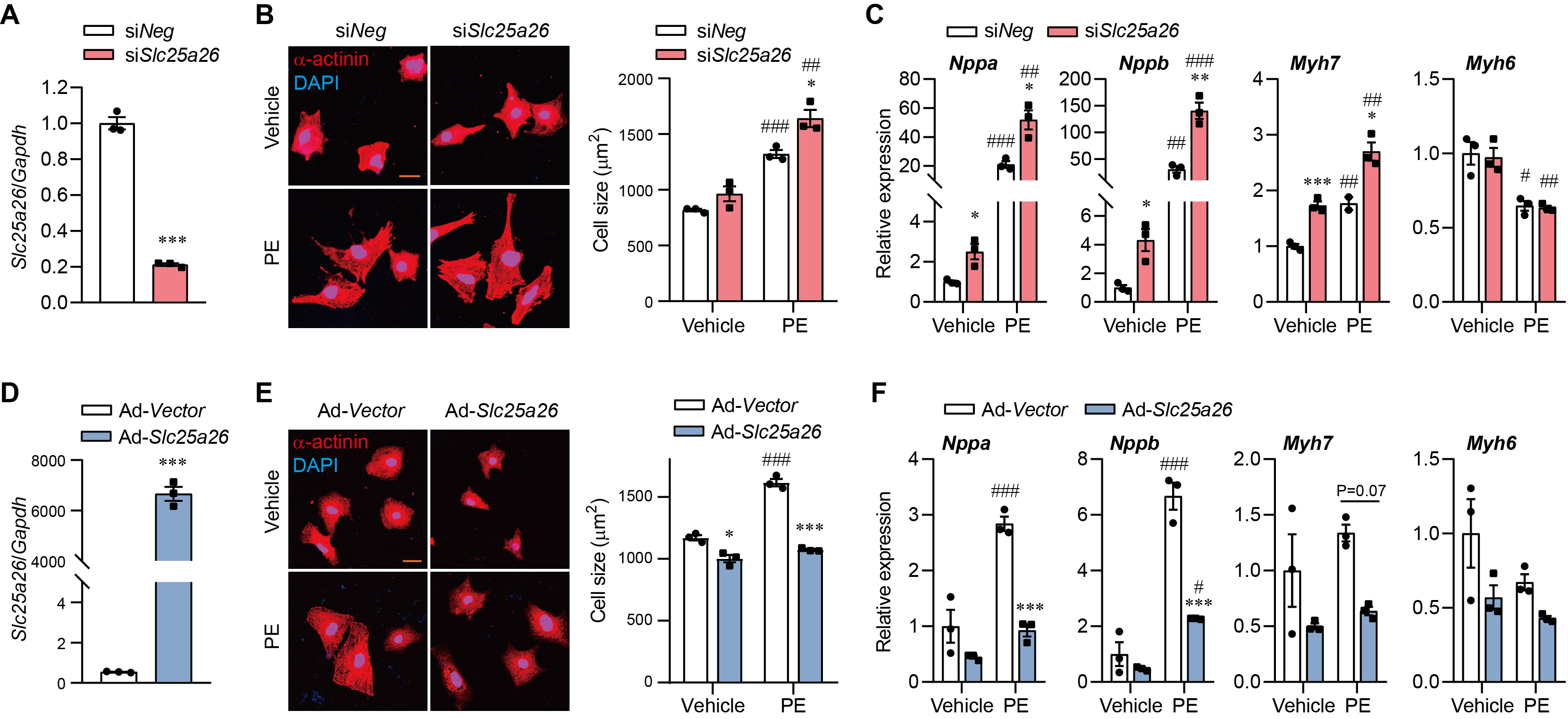

### Figure S3. SLC25A26 mitigates phenylephrine (PE)-induced hypertrophy in neonatal rat ventricular myocytes (NRVMs).

**A**, Validation of *Slc25a26* knockdown by a specific siRNA (siSlc25a26) in NRVMs measured by qRT-PCR. siNeg, negative control siRNA; *N* = 3.

**B**, Representative α-actinin staining images (left) and WGA staining quantification (right) showing the impact of *Slc25a26* knockdown on PE-induced cell size enlargement in NRVMs. Scale bar, 10μm. *N* = 3.

**C**, Impact of *Slc25a26* knockdown on the expressions of *Nppa*, *Nppb*, *Myh7*, and *Myh6* relative to the expression of *Gapdh* in PE-treated NRVMs. *N* = 3.

**D**, Validation of *Slc25a26* overexpression in NRVMs infected by a constructed adenovirus (Ad-Slc25a26) with Ad-Vector. *N* = 3.

**E**, Representative α-actinin staining images (left) and WGA staining quantification (right) showing the impact of *Slc25a26* overexpression on PE-induced cell size enlargement in NRVMs. Scale bar, 10μm. *N* = 3.

**F**, Impact of *Slc25a26* overexpression on the expressions of *Nppa*, *Nppb*, *Myh7*, and *Myh6* relative to the expression of *Gapdh* in PE-treated NRVMs. *N* = 3.

Data were presented as mean ± SEM of at least three independent experiments, unless denoted elsewhere. *p < 0.05, **p < 0.01, ***p < 0.001 vs. si*Neg* or Ad-*vector*; ^#^*P* < 0.05, ^##^*P* < 0.01, ^###^*P* < 0.001 vs. vehicle, (**A** and **D**, unpaired Student’s t test; **B**, **C**, **E** and **F**, two-way ANOVA with Tukey’s multiple comparison test).

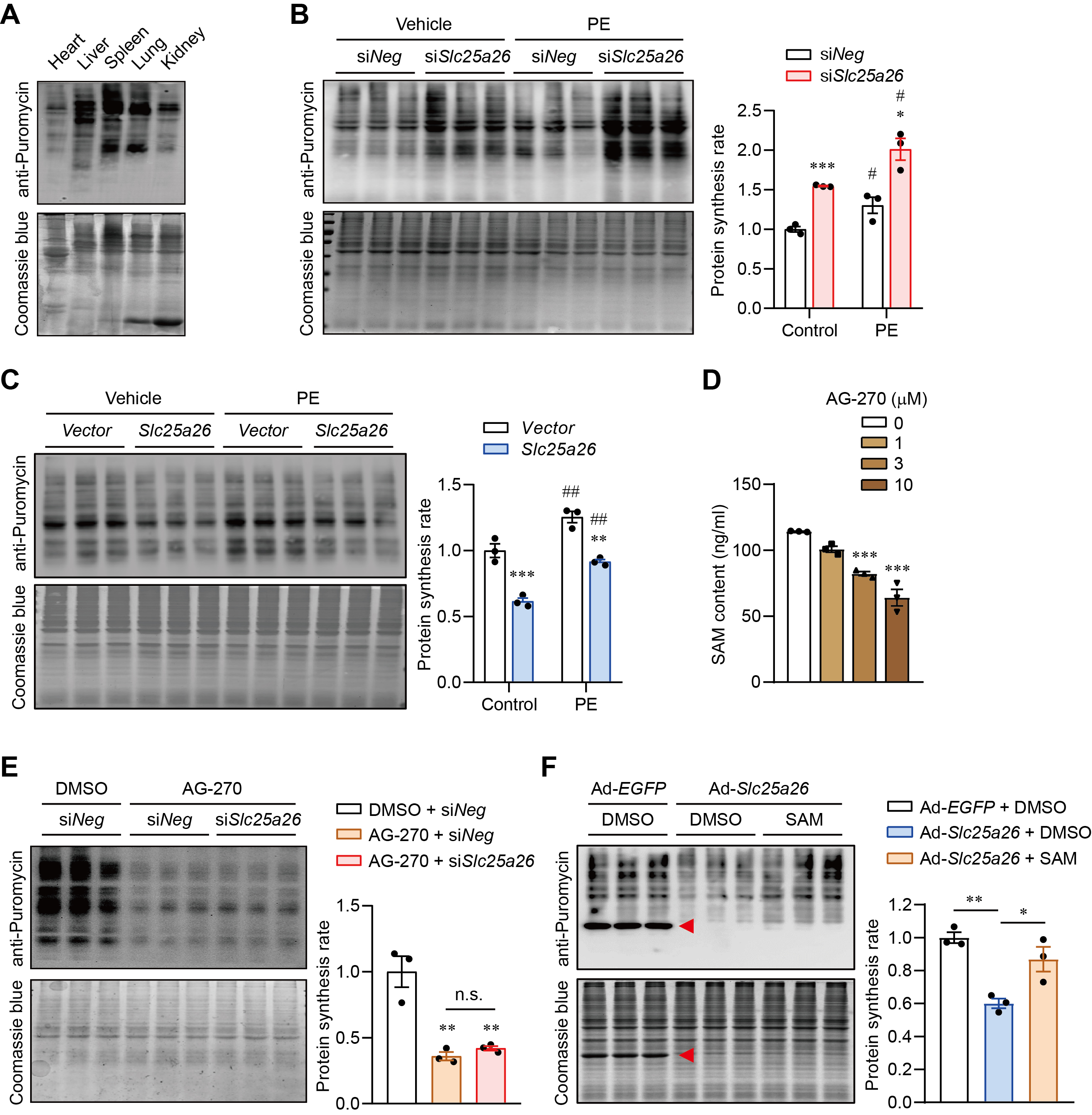

### Figure S4. SLC25A26 regulates translation through redistributing SAM.

**A**, Puromycin incorporation immunoblot using anti-puromycin antibody showing the protein synthesis rate in different mouse tissues. Coomassie blue staining was used as the loading control.

**B**, Puromycin incorporation immunoblot (left) and quantification (right) showing the protein synthesis rate (normalized to Coomassie blue staining) in NRVMs with and without PE treatment or *Slc25a26* knockdown. *N* = 3.

**C**, Protein synthesis rate in NRVMs with and without PE treatment or *Slc25a26* overexpression. *Slc25a26*-expressing or vector plasmids were transfected into cells using nanoparticle reagents. *N* = 3.

**D**, Dose-dependent effects of the MAT inhibitor AG-270 on the SAM content in NRVMs measured by an enzyme-linked immunosorbent assay (ELISA) kit. *N* = 3.

**E**, Impact of AG-270 (3μM) on protein synthesis rate in NRVMs with and without *Slc25a26* knockdown. *N* = 3.

**F**, Impact of SAM supplementation (1μM) on protein synthesis rate in NRVMs with adenovirus-mediated *Slc25a26* overexpression. *N* = 3.

Data were presented as mean ± SEM of at least three independent experiments, unless denoted elsewhere. n.s., not significant, *p < 0.05, **p < 0.01, ***p < 0.001 vs. si*Neg* or *vector* or DMSO+si*Neg* or Ad-*EGFP*+DMSO; ^#^*P* < 0.05, ^##^*P* < 0.01, ^###^*P* < 0.001 vs. vehicle, (**D**, **E** and **F**, one-way ANOVA with Tukey’s multiple comparison test; **B** and **C**, two-way ANOVA with Tukey’s multiple comparison test).

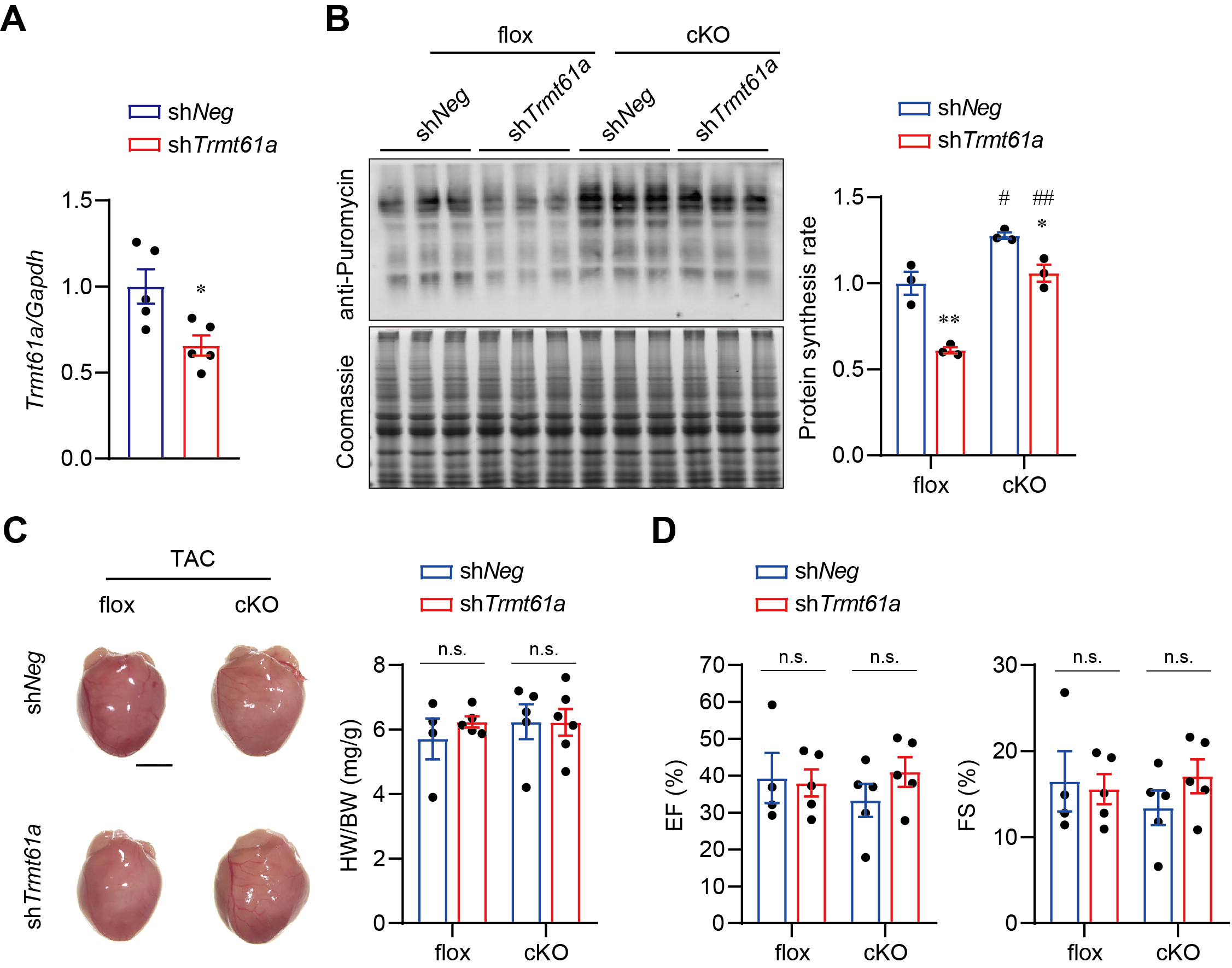

### Figure S5. Silencing *Trmt61a in vivo* alleviates the acceleration of protein synthesis in *Slc25a26*-cKO hearts, but does not rescue cardiac hypertrophy.

**A**, Validation of the *Trmt61a* knockdown efficiency in mice infected with adeno-associated viruses expressing shTrmt61a or a negative control shRNA (shNeg). *N* = 5.

**B**, Immunoblots (left) and quantification (right) showing the impact of *Trmt61a* knockdown on protein synthesis rate in *Slc25a26*-flox and *Slc25a26*-cKO mice. *N* = 3.

**C**, Representative morphology images (left) and weights (right) of the *Slc25a26*-flox and *Slc25a26*-cKO hearts showing the impact of *Trmt61a* knockdown on TAC-induced cardiac hypertrophy. Scale bar, 3mm; *N* = 4~6 as indicated by dots on each bar.

**D**, Impact of *Trmt61a* knockdown on post-TAC cardiac ejection fraction (EF) and fraction shortening (FS) of the *Slc25a26*-flox and *Slc25a26*-cKO hearts. *N* = 4~5 as indicated by dots on each bar.

Data were presented as mean ± SEM of at least three independent experiments, unless denoted elsewhere. n.s., not significant, *p < 0.05, **p < 0.01, ***p < 0.001 vs. sh*Neg*; ^#^*P* < 0.05, ^##^*P* < 0.01, ^###^*P* < 0.001 vs. flox, (**A**, unpaired Student’s t test; **B**, **C** and **D**, two-way ANOVA with Tukey’s multiple comparison test).

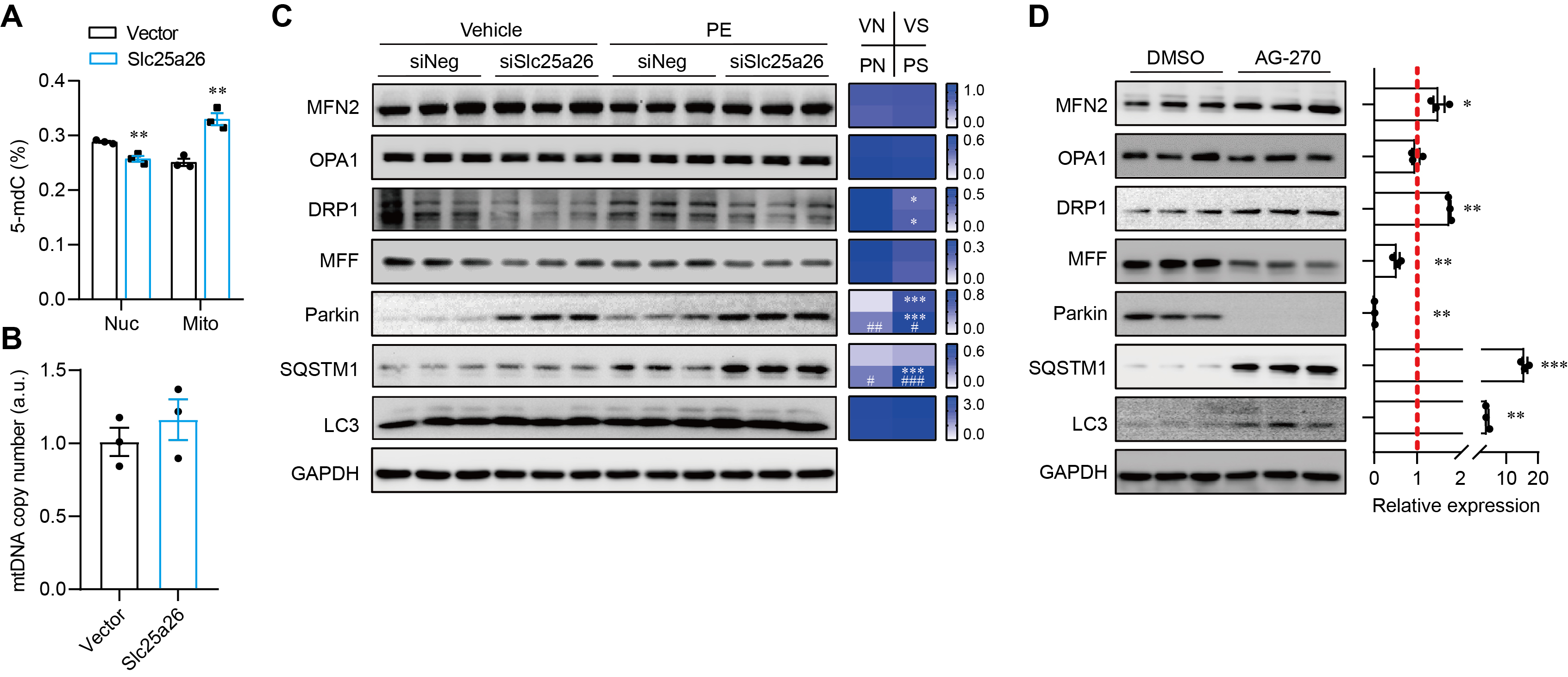

### Figure S6. SLC25A26-mediated mitoSAM accumulation promotes mitochondrial quality control.

**A**, Impact of *Slc25a26* overexpression on nuclear (Nuc) and mitochondrial (Mito) DNA 5-mdC modification in NRVMs measured by ELISA. *N* = 3.

**B**, Impact of *Slc25a26* overexpression on mtDNA copy number (Cytb/H19) in NRVMs measured by qRT-PCR. *N* = 3.

**C** and **D**, Immunoblots (left) and quantification (right) of proteins involved in mitochondrial dynamics in PE-treated NRVMs with *Slc25a26* knockdown (**C**) and in NRVMs treated with AG-270 (**D**). *N* = 3.

Data were presented as mean ± SEM of at least three independent experiments, unless denoted elsewhere. *p < 0.05, **p < 0.01, ***p < 0.001 vs. vector; ^#^*P* < 0.05, ^##^*P* < 0.01, ^###^*P* < 0.001 vs. vehicle, (**A**, **B** and **D** unpaired Student’s t test; **C**, two-way ANOVA with Tukey’s multiple comparison test).
